# Casirivimab and imdevimab for the treatment of hospitalized patients with COVID-19

**DOI:** 10.1101/2021.11.05.21265656

**Authors:** Selin Somersan-Karakaya, Eleftherios Mylonakis, Vidya P. Menon, Jason C. Wells, Shazia Ali, Sumathi Sivapalasingam, Yiping Sun, Rafia Bhore, Jingning Mei, Jutta Miller, Lisa Cupelli, Andrea T. Hooper, Jennifer D. Hamilton, Cynthia Pan, Viet Pham, Yuming Zhao, Romana Hosain, Adnan Mahmood, John D. Davis, Kenneth C. Turner, Yunji Kim, Amanda Cook, Bari Kowal, Yuhwen Soo, A. Thomas DiCioccio, Gregory P. Geba, Neil Stahl, Leah Lipsich, Ned Braunstein, Gary A. Herman, George D. Yancopoulos, David M. Weinreich, COVID-19 Phase 2/3 Hospitalized Trial Team

**Affiliations:** Regeneron Pharmaceuticals, Inc., Tarrytown, NY, USA; Brown University, Providence, RI, USA; NYC Health + Hospitals/Lincoln, The Bronx, NY, USA; The Oregon Clinic, Portland, OR, USA

**Author notes:** **Correspondence to:** Selin Somersan-Karakaya, MD,. S. S-K. and E. M. contributed equally. Former employee of Regeneron Pharmaceuticals, Inc (Sumathi Sivapalasingam, Leah Lipsich).

## Abstract

**Background:** Hospitalized patients with COVID-19 experience high mortality rates, ranging from 10% to 30%. Combined casirivimab and imdevimab (CAS+IMD) is authorized for use in outpatients with COVID-19 and in post-exposure prophylaxis. The UK-based platform RECOVERY study reported improved survival in hospitalized seronegative patients treated with CAS+IMD; however, in most of the world, anti-spike monoclonal antibody therapy is currently not approved for hospitalized patients.

**Methods:** In this phase I/II/III double-blind placebo-controlled trial, patients hospitalized with COVID-19 were randomized (1:1:1) to 2.4 g or 8.0 g of CAS+IMD or placebo, and characterized at baseline for viral load and SARS-CoV-2 endogenous immune response.

**Results:** 1336 patients on low-flow or no supplemental oxygen were treated. The primary endpoint was met: in seronegative patients, the least squares mean difference (CAS+IMD vs placebo) for time-weighted average change from baseline viral load was –0.28 log_10_ copies/mL (95% confidence interval [CI] –0.51 to –0.05; *P* = .0172). The primary clinical analysis of death or mechanical ventilation from day 6 to 29 in patients with high viral load had a strong positive trend but did not reach significance. CAS+IMD numerically reduced all-cause mortality in seronegative patients through day 29 (relative risk reduction, 55.6%; 95% CI 24.2–74.0; nominal *P* = .0032). No safety concerns were noted.

**Conclusions:** In hospitalized patients with COVID-19 on low-flow or no oxygen, CAS+IMD treatment reduced viral load and the risk of death or mechanical ventilation as well as all-cause mortality in the overall population, with the benefit driven by seronegative patients and no harm observed in seropositive patients.

## Introduction

The clinical progression of coronavirus disease 2019 (COVID-19), caused by severe acute respiratory syndrome coronavirus 2 (SARS-CoV-2), is highly variable; while many cases manifest with relatively mild symptoms, others progress to severe respiratory failure requiring supplemental oxygen and/or mechanical ventilation [1-4]. Casirivimab and imdevimab (CAS+IMD) is a monoclonal antibody combination approved or authorized for emergency use for the treatment of outpatients with mild-to-moderate COVID-19, and for post-exposure prophylaxis in the United States and other jurisdictions [5-7]. In outpatients with COVID-19, CAS+IMD reduced hospitalization or all-cause death, reduced viral load, and shortened symptom duration [8-10]. Data show that CAS+IMD is also highly effective in preventing asymptomatic as well as symptomatic COVID-19, evidenced by a single-dose subcutaneous administration showing ∼80% lower risk of developing COVID-19 for household contacts living with an infected individual [11]. The totality of evidence suggests that benefit is greatest when treated early [12].

Based on the potent anti-viral activity of CAS+IMD, it was prospectively hypothesized that reducing viral burden as early as possible would also decrease morbidity and mortality associated with SARS-CoV-2 infection in hospitalized patients. In a recent open-label platform trial of hospitalized patients with COVID-19 in the United Kingdom (RECOVERY), CAS+IMD met its primary endpoint in improving overall survival in patients who had not mounted their own immune response at baseline (seronegative) by 20%, and also improved duration of hospitalization [13]. Here, we describe the final efficacy and safety results from the first phase I/II/III double-blind placebo-controlled trial of CAS+IMD in hospitalized patients with COVID-19 on low-flow or no supplemental oxygen.

## Methods

### Trial Design

This was an adaptive, phase I/II/III, double-blinded, placebo-controlled trial to evaluate the efficacy, safety, and tolerability of CAS+IMD in hospitalized adult patients with COVID-19, conducted at 103 sites in the United States, Brazil, Chile, Mexico, Moldova, and Romania (NCT04426695).

Patients were enrolled in 1 of 4 cohorts based on disease severity: no supplemental oxygen (cohort 1A), low-flow oxygen (cohort 1), high-intensity oxygen (cohort 2), or mechanical ventilation (cohort 3), as described in the patient cohorts section in the appendix (**Supplementary Figure 1**). The trial proceeded through phase II for patients requiring no supplemental oxygen (cohort 1A) and phase III for patients requiring low-flow oxygen (cohort 1); together, these patients are the subject of this manuscript. As phase I/II data from patients on low-flow oxygen was previously unblinded in an interim analysis on December 22, 2020, it was not included in the phase III efficacy analyses. For patients requiring high-intensity oxygen (cohort 2) or mechanical ventilation (cohort 3), enrollment was paused early in the study based on recommendation of the independent data monitoring committee (IDMC) and data were not included due to low sample size; this is further described in the trial adaptations section of the appendix.

Patients were randomized 1:1:1 to a single intravenous dose of 2.4 g CAS+IMD (1.2 g casirivimab and 1.2 g imdevimab), 8.0 g CAS+IMD (4.0 g casirivimab and 4.0 g imdevimab), or placebo. Within each cohort, randomization was stratified by standard-of-care treatment (antiviral therapies, non-antiviral therapies; phase I/II/III) and country (phase II/III). The trial included a screening/baseline period (days –1 to 1), a hospitalization/post-discharge period, a monthly follow-up period, and an end-of-study visit (phase I day 169, phase II/III day 57; **Supplementary Figure 1**).

### Patients

Patients were 18 years of age or older and hospitalized with confirmed SARS-CoV-2 within 72 h, with symptom onset ≤10 days from randomization. Standard-of-care treatments for COVID-19, per the investigator, were permitted. Full inclusion and exclusion criteria are presented in the appendix.

### Outcome Measures

The primary virologic efficacy endpoint was the time-weighted average (TWA) daily change from baseline (day 1) viral load in nasopharyngeal samples through day 7 in the seronegative population [10]. The primary clinical efficacy endpoint was the proportion of patients who died or required mechanical ventilation from days 6 to 29 and days 1 to 29 for the high-viral load, seronegative, and overall populations, tested in a statistical hierarchy (**Supplementary Table 1**). Clinical efficacy from days 6 to 29 was included as part of the hierarchical testing strategy because several days of viral suppression in this severe population may be required before clinical impact is observed. High viral load was selected for the first clinical efficacy endpoint in the hierarchy based on previous experience with treatment in the outpatient setting [8, 10]; it was expected that the high viral load population would be highly correlated with the seronegative population, and assessing viral load could be easier than assessing serostatus in a clinical setting.

Secondary efficacy endpoints examined all-cause mortality and discharge from/readmission to hospital. Safety endpoints included the proportion of patients with treatment-emergent serious adverse events (SAEs) and adverse events of special interest (AESIs): infusion-related reactions (IRRs) through day 4, and grade ≥2 hypersensitivity reactions through day 29.

### Statistical Analysis

The statistical analysis plan was finalized prior to database lock and unblinding. The full analysis set (FAS) was used for safety analyses and includes all randomized patients who received any amount of study drug. The modified FAS (mFAS) was used for efficacy analyses and excludes patients who had negative central lab SARS-CoV-2 quantitative reverse transcriptase polymerase chain reaction at baseline.

The primary virologic endpoint was analyzed using the analysis of covariance model, and primary clinical endpoints were analyzed using either the exact method for binomial distribution or asymptotic normal approximation method, as predefined in the statistical analysis plan; details are provided in the appendix. As the trial was stopped earlier than planned (due to low enrolment prior to the surge associated with the delta variant), the sample size was smaller than anticipated and it was elected to combine the CAS+IMD dose groups and pool patients on no supplemental oxygen (phase II) and low-flow oxygen (phase III) for efficacy measures. The multiplicity adjustment approach, a hierarchical procedure, was used to control the overall type-1 error rate at 0.05 for the primary virologic and clinical outcome endpoints (**Supplementary Table 1**). Where an endpoint in the hierarchy did not reach statistical significance, all *P* values for subsequent comparisons were nominal; other analyses, including all-cause mortality, were reported descriptively.

Safety was assessed in separate analyses for patients receiving no supplemental oxygen (phase II) and low-flow oxygen (phase I/II/III). Prespecified subgroup analyses using baseline serostatus and viral load were selected based on previous results [10]. Sample size calculations and missing data handling are described in the appendix.

## Results

### Demographics and Baseline Characteristics

As of April 9, 2021, 1364 patients on low-flow or no supplemental oxygen were randomized into the study, of whom 1336 were treated. Of those, 1197 (89.6%) tested positive centrally for SARS-CoV-2 (constituting the mFAS) with 406, 398, and 393 in the CAS+IMD 2.4 g, 8.0 g, and placebo groups, respectively (**Supplementary Figure 2**).

Baseline demographics were well-balanced. The median age was 62 years, 54.1% were male, mean body mass index was 31.1 kg/m^2^, 12.1% identified as Black/African American, and 30.1% identified as Hispanic/Latino (**Table 1**). COVID-19 characteristics were similar except for a higher proportion of seropositive patients in the placebo group (51.1%) compared to the combined CAS+IMD group (45.9%; **Table 1**). The number of days of COVID-19 symptoms, viral load, and C-reactive protein concentrations at baseline were balanced, and similar proportions of patients received remdesivir and/or systemic corticosteroids (**Table 1**). Demographics and baseline characteristics by serostatus are presented in **Supplementary Table 2**.

**Table 1.**
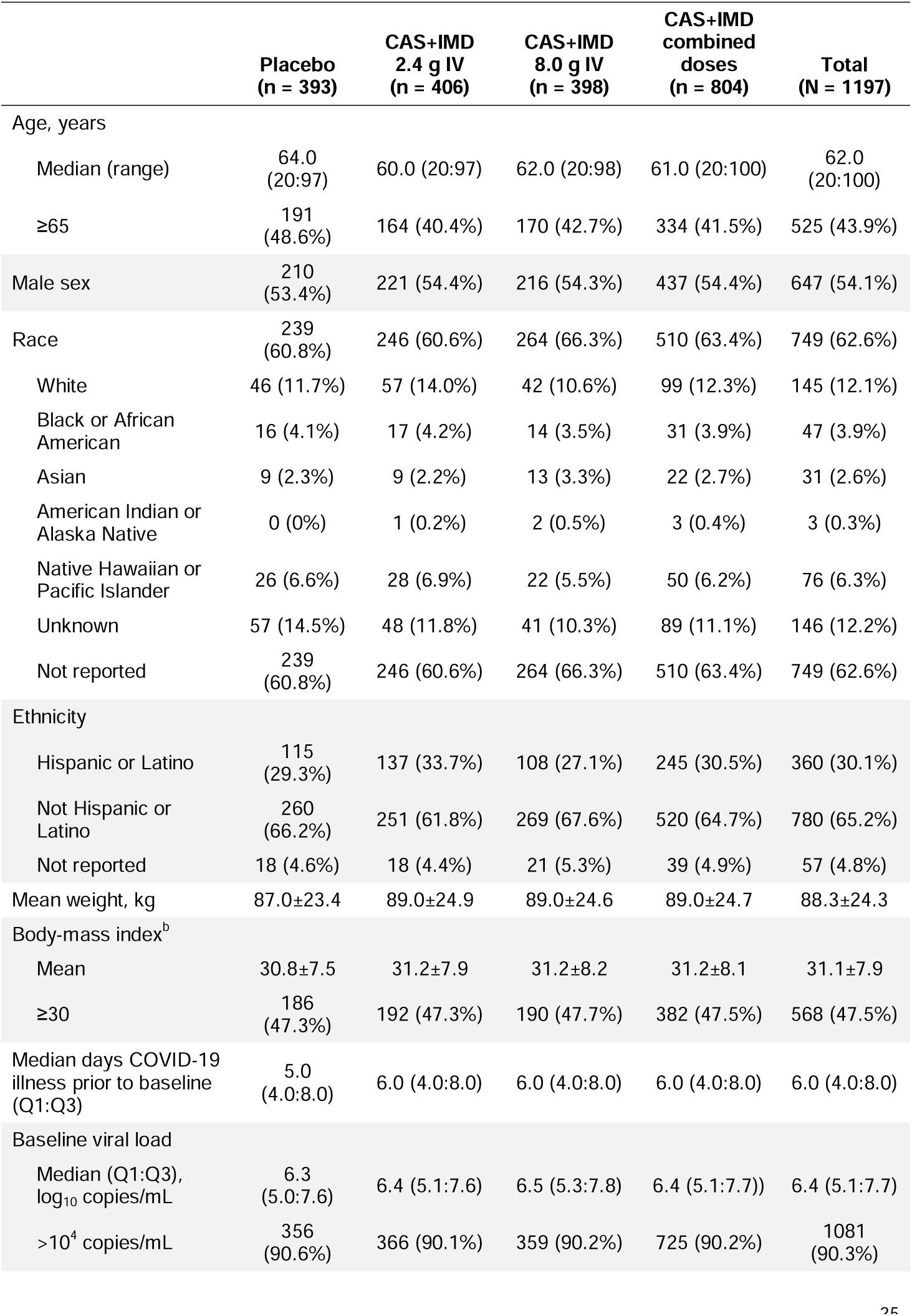
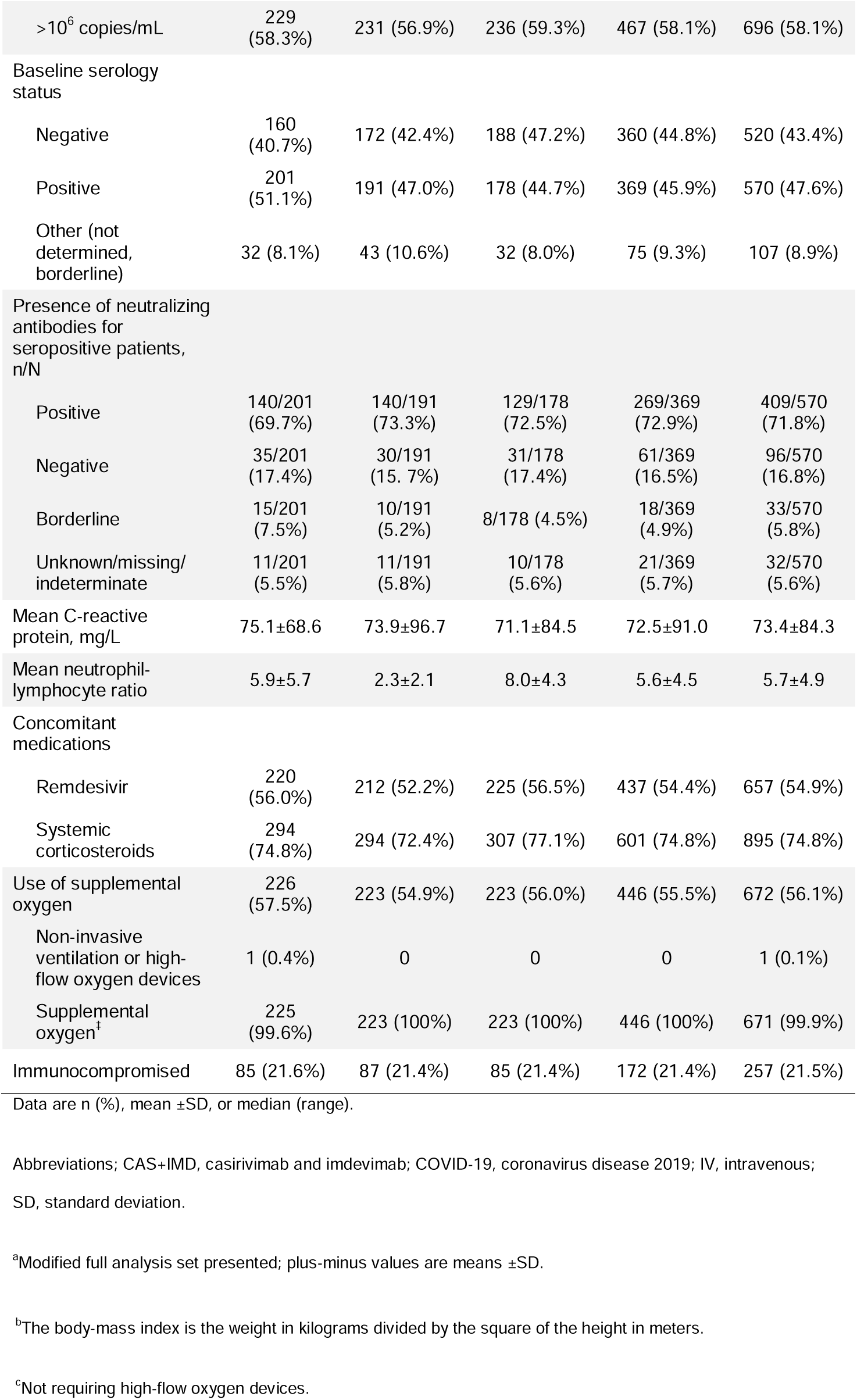
Demographics and baseline characteristics^a^.

### Virologic Efficacy

CAS+IMD significantly reduced viral load in seronegative patients on low-flow or no supplemental oxygen; the least-squares (LS) mean (95% confidence interval [CI]) TWA daily change in viral load from baseline through day 7 was –1.03 log_10_ copies/mL (95% CI –1.22 to –0.84) in the placebo group compared with –1.31 log_10_ copies/mL (95% CI –1.43 to –1.18) in the CAS+IMD combined dose group, with an LS mean difference versus placebo of –0.28 log_10_ copies/mL (95% CI –0.51 to –0.05; *P* = .0172; **Supplementary Table 2**).

Both doses of CAS+IMD exhibited similar viral load reductions, showing improvement over placebo starting at day 3 and reaching significance at day 7, after which viral load in the CAS+IMD groups continued to fall relative to placebo (**Figure 1, Supplementary Figure 3**). The maximum LS mean differences versus placebo in seronegative patients were at day 7 (described above), day 9 (–0.47 log_10_ copies/mL, 95% CI –0.71 to –0.23), and day 11 (–0.59 log_10_ copies/mL, 95% CI – 0.85 to –0.34; **Figure 1A**). The overall population LS mean fell below the lower limit of quantification (2.85 log_10_ copies/mL) 2 days earlier with CAS+IMD (day 9 CAS+IMD vs day 11 placebo; **Supplementary Figure 3**). Reductions of viral load were observed in the seronegative (**Figure 1A**) and the overall populations (**Supplementary Figure 3**), with greater reductions in seronegative patients.

**Figure 1.**
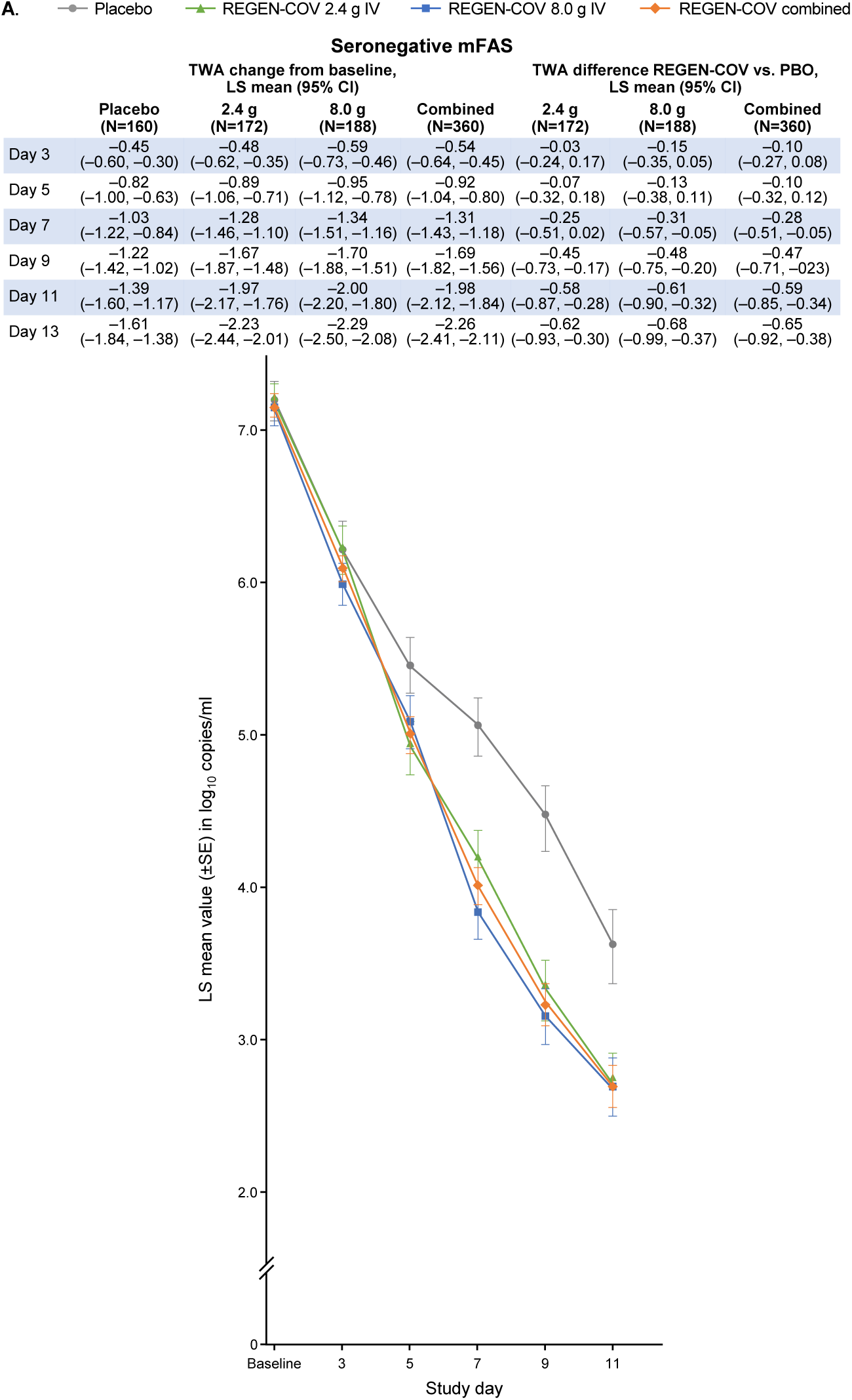
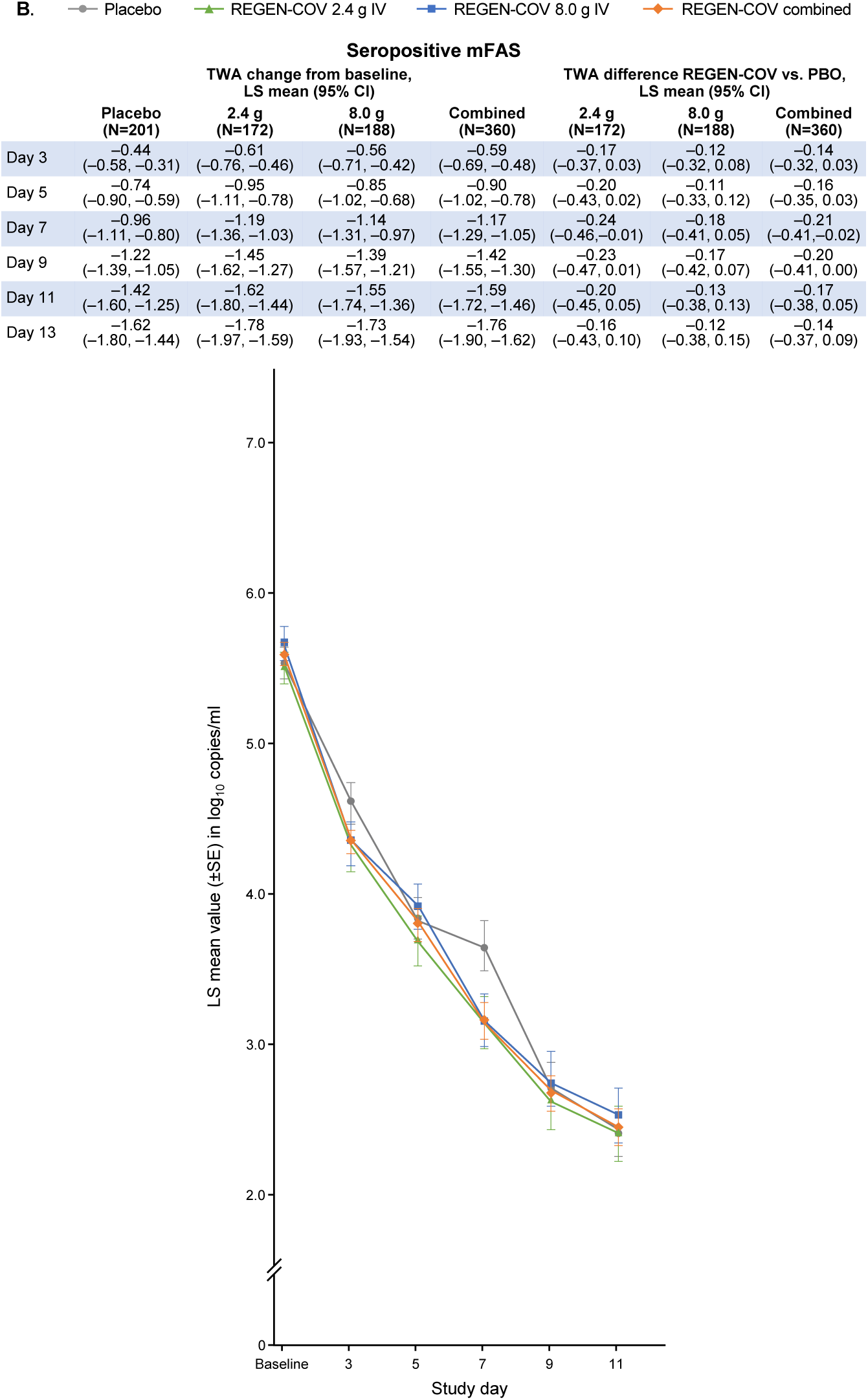
Viral Load by Serostatus. Panel A graph shows LS mean viral load following administration of CAS+IMD (2.4 g, 8.0 g, or combined analysis of 2.4 and 8.0 g) or placebo for patients who tested negative for all SARS-CoV-2 antibodies at baseline (seronegative). Panel B shows the same but for patents who tested positive for any SARS-CoV-2 antibody at baseline (seropositive). For both panels, the lower limit of quantification is 2.85 log_10_ copies/mL. Abbreviations: CAS+IMD, casirivimab and imdevimab; CI, confidence interval; IV, intravenous; mFAS, modified full analysis set; LS, least-squares; PBO, placebo; SARS-CoV-2, severe acute respiratory syndrome coronavirus 2; SE, standard error; TWA, time-weighted average.

### Clinical Efficacy

#### Death or Mechanical Ventilation

Endpoints were examined both from days 1 to 29 and days 6 to 29, and were evaluated in the seronegative, high-viral load, and overall populations, as described in the Methods. The analyses presented herein examine the pooled CAS+IMD dose group and pooled cohorts for low-flow and no supplemental oxygen (**Figure 2**). Individual dose groups of 2.4 g and 8.0 g of CAS+IMD (**Supplementary Figure S4**) and separate cohorts by respiratory status (**Supplementary Figure S5**) were also examined, and showed trends of benefit in seronegative patients across all clinical endpoints.

**Figure 2.**
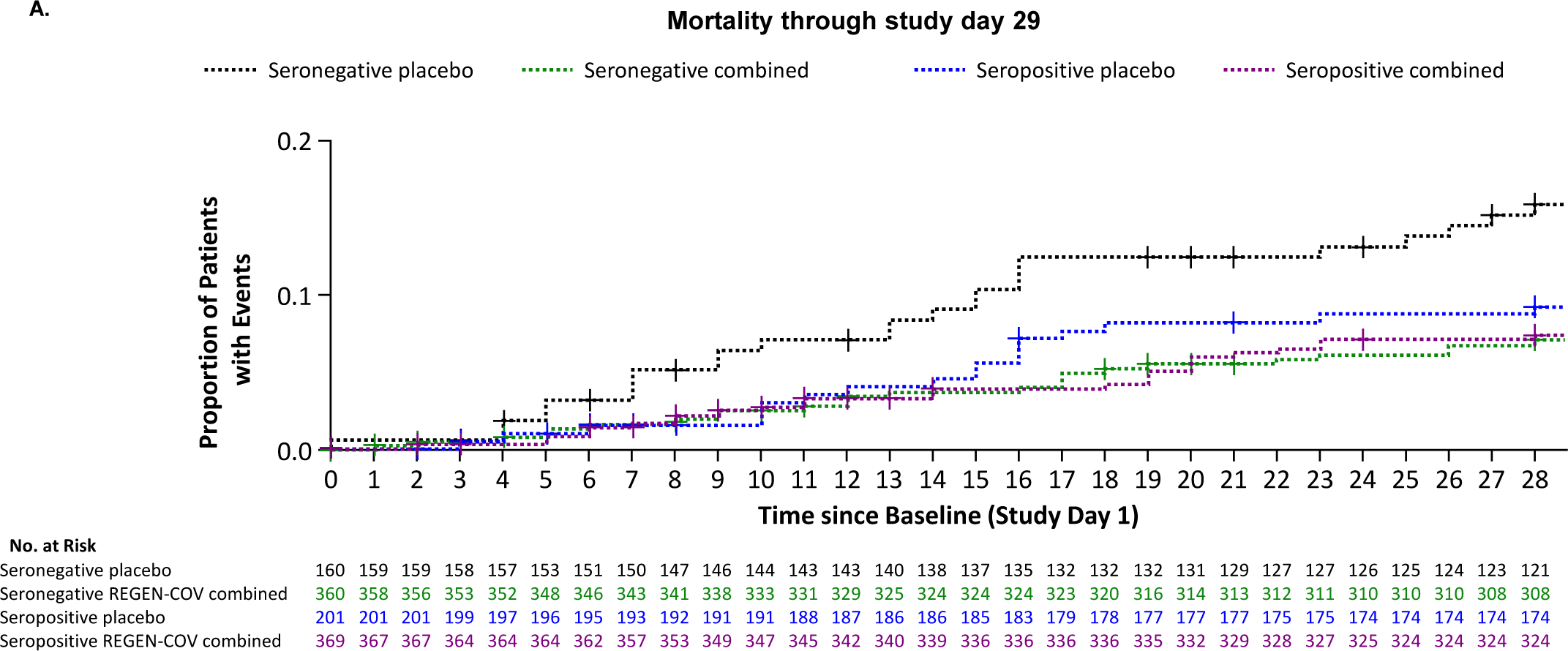
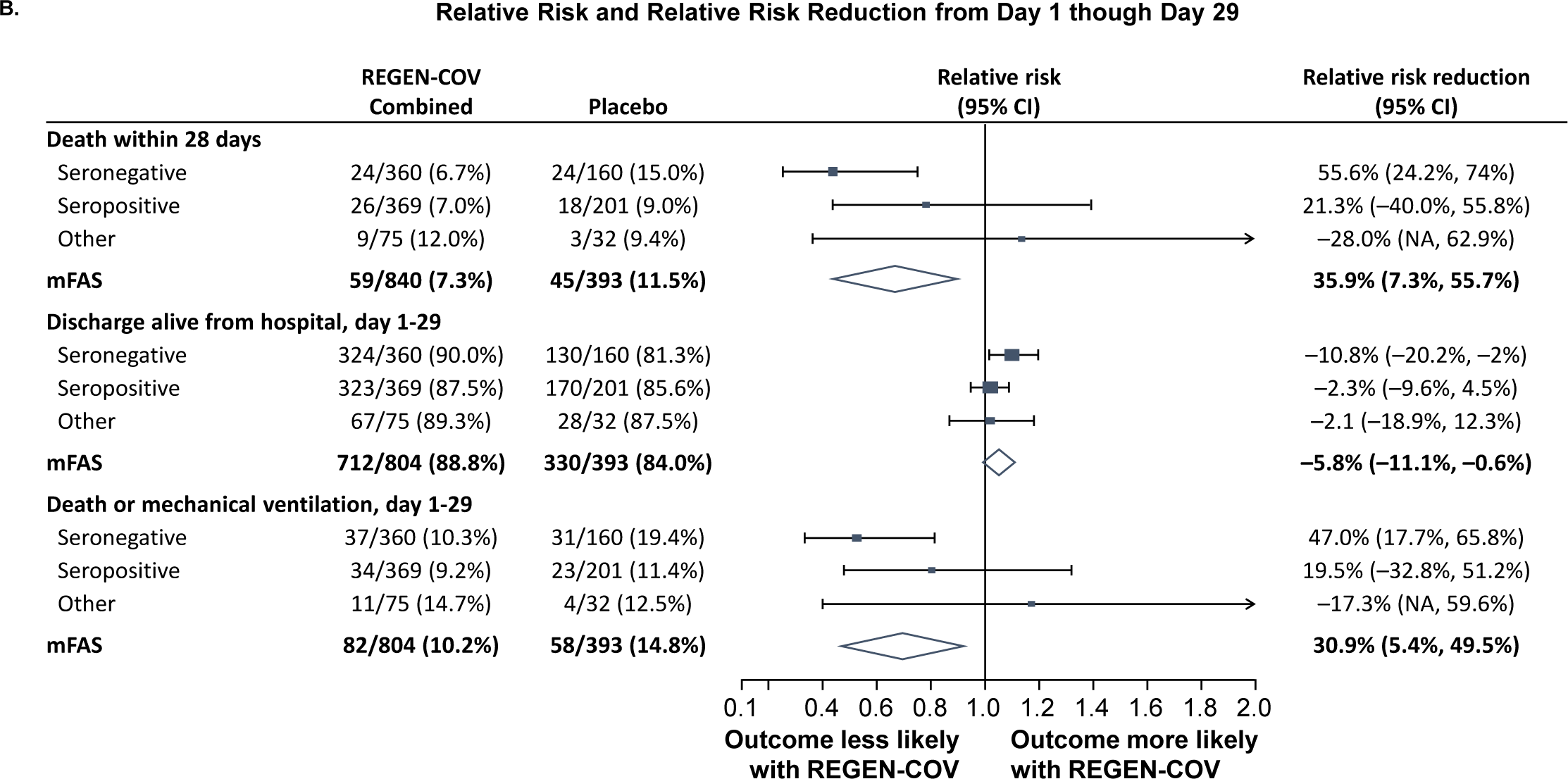
Efficacy Outcomes by Serostatus for Combined Dose CAS+IMD from Day 1 though Day 29. Panel A shows a Kaplan–Meier curve for the proportion of patients who died through study day 29, after administration of CAS+IMD (combined analysis of 2.4 g or 8.0 g) or placebo. Results are analyzed separately for patients who were seronegative or seropositive at baseline. + indicates censoring. Panel B shows a forest plot of relative risk and relative risk reduction with 95% CIs for CAS+IMD combined dose analysis (2.4 g and 8.0 g) versus placebo. Parameters examined include death within 28 days, discharge alive from hospital from days 1 to 29, and death or mechanical ventilation from days 1 to 29. For all populations, the mFAS was comprised of patients who tested positive for SARS-CoV-2 at baseline. Populations analyzed include patients who tested negative for all SARS-CoV-2 antibodies at baseline (seronegative mFAS), patients who tested positive for any SARS-CoV-2 antibody at baseline (seropositive mFAS), those with inconclusive or missing baseline serology (other), and the overall population regardless of serostatus (overall mFAS). For the proportion of death within 28 days and the proportion of death or mechanical ventilation with 28 days, the lower bounds of the CI of the relative risk reduction were –342.0% and –241.0%, respectively, which are presented as “NA” in the figure. *P* values are considered nominal. Abbreviations: CAS+IMD, casirivimab and imdevimab; CI, confidence interval; mFAS, modified full analysis set; SARS-CoV-2, severe acute respiratory syndrome coronavirus 2.

In the statistical hierarchy (**Supplementary Table 1**), the first test for clinical efficacy on the endpoint of death or mechanical ventilation in the high viral load population from days 6 to 29 showed a numerically lower risk compared to placebo but did not reach statistical significance (relative risk reduction [RRR], 25.5%; 95% CI –16.2–52.2; *P* = .2048; **Table 2**); accordingly, all subsequent clinical efficacy analyses are considered descriptive, with nominal *P* values. The second test for clinical efficacy on the endpoint of death or mechanical ventilation in the seronegative population from days 6 to 29 showed an RRR of 47.1% (95% CI 10.2– 68.8; nominal *P* = .0195 **Table 2**). Trends of improvement were also observed from days 6 to 29 in the overall population (RRR, 24.2%; 95% CI –10.9–48.2; nominal *P* = .1486; **Table 2**).

**Table 2.**
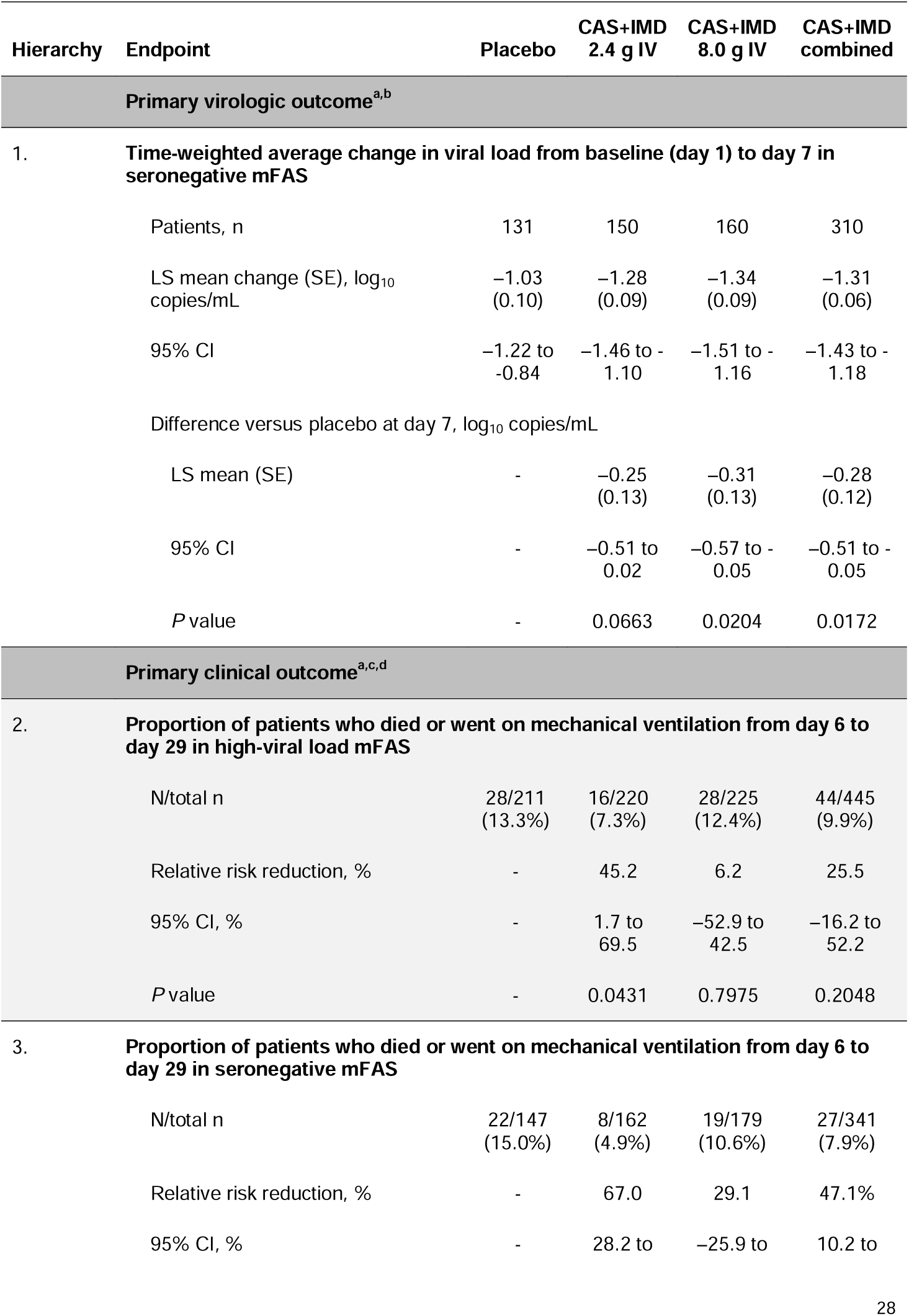
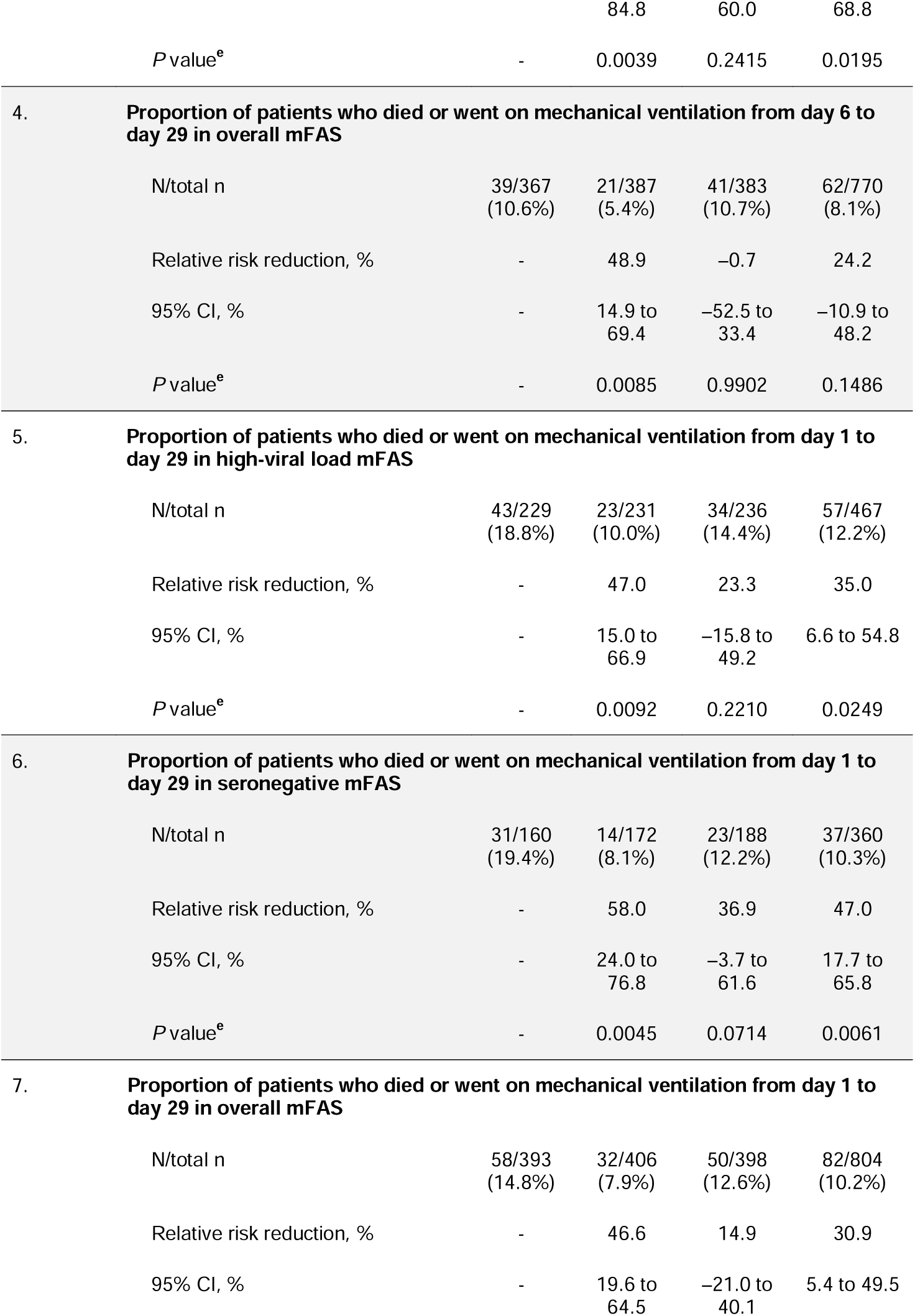
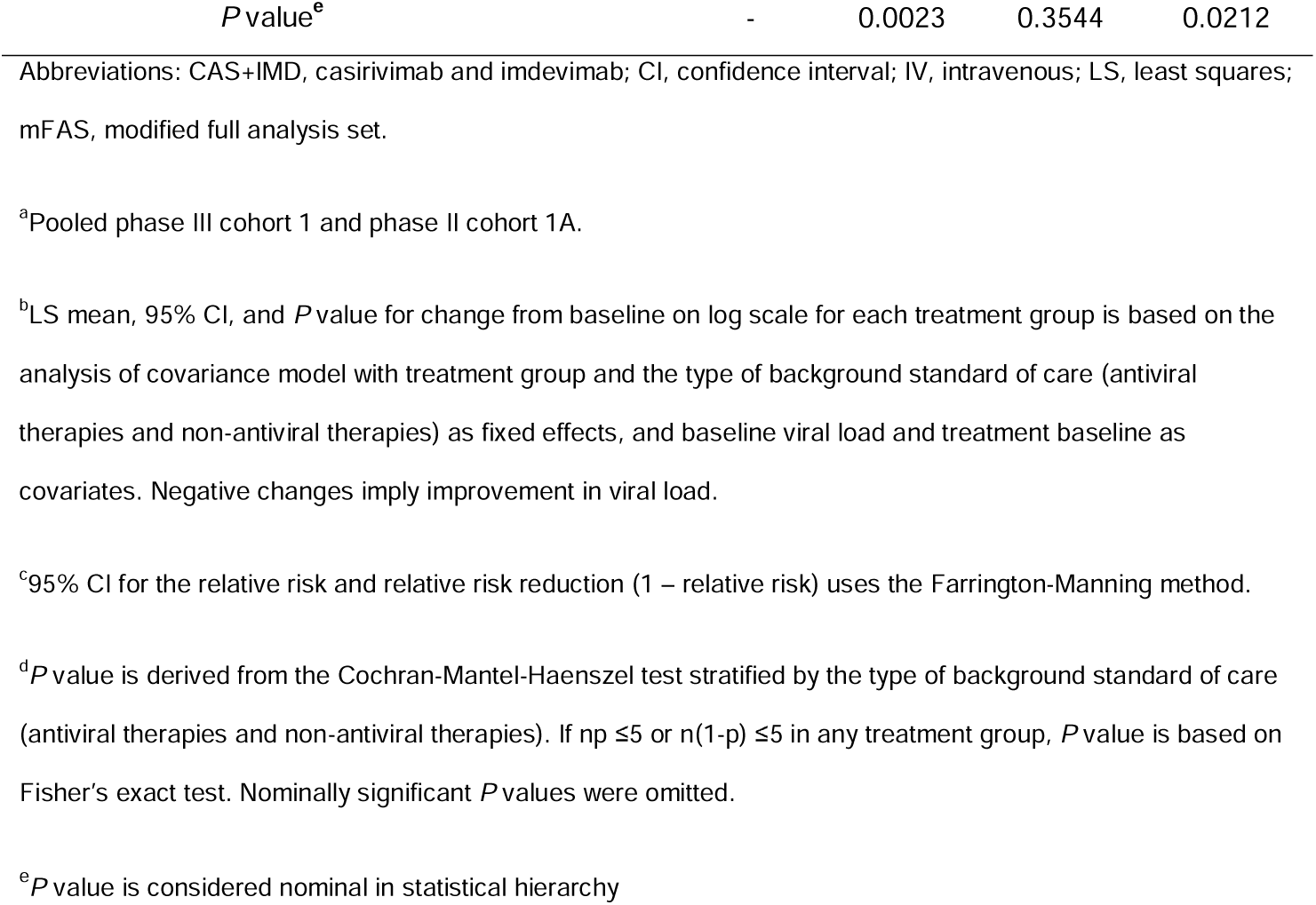
Primary virologic and clinical endpoints.

Treatment with CAS+IMD led to a reduction in the proportions of patients who died or required mechanical ventilation, with improvement from days 1 to 29 in the high viral load (RRR, 35.0%; 95% CI 6.6–54.8, nominal *P* = .0249), seronegative (RRR, 47.0%; 95% CI 17.7–65.8; nominal *P* = .0061), and overall (RRR, 30.9%; 95% CI 5.4–49.5; nominal *P* = .0212) populations (**Table 2**). While seronegative patients exhibited the greatest benefit from CAS+IMD treatment, no meaningful benefit or harm was observed in seropositive patients (RRR, 19.5%; 95% CI –32.8–51.2; nominal *P* = .3010; **Figure 2**).

#### All-Cause Mortality

Treatment with CAS+IMD led to numeric improvement in all-cause mortality through day 29 in the seronegative, high-viral load, and overall populations. The greatest reduction in the relative risk of death occurred in seronegative patients; 24/360 (6.7%) died within 28 days in the CAS+IMD group, compared to 24/160 (15.0%) in the placebo group (RRR, 55.6%; 95% CI 24.2–74.0; nominal *P* = .0032; **Figure 2B**). No harm or meaningful benefit was observed in the seropositive population (**Figure 2A**). For the overall population, driven by the seronegative group, a substantial reduction in death was observed in which 59/804 patients (7.3%) died within 28 days in the CAS+IMD combined dose group, compared to 45/393 patients (11.5%) in the placebo group (RRR, 35.9%; 95% CI 7.3–55.7; nominal *P* = .0178; **Figure 2B**). The improvement in all-cause mortality with CAS+IMD persisted through study day 57 (**Supplementary Figure 6**). Similar results were also observed in the secondary endpoints of hospital discharge and readmission, shown in **Supplementary Table 3** and **Supplementary Table 4**, and described in the appendix.

### Safety

SAEs were experienced by more patients in the placebo group than the CAS+IMD group for patients on low-flow oxygen (131/469 [27.9%] placebo vs 224/941 [23.8%] CAS+IMD) and no supplemental oxygen (43/198 [21.7%] placebo vs 61/399 [15.3%] CAS+IMD; **Table 3**). More patients experienced treatment-emergent adverse events that resulted in death in the placebo group compared with CAS+IMD for patients on low-flow oxygen (72/469 [15.4%] placebo vs 108/941 [11.5%] CAS+IMD **Supplementary Table 5**) and no supplemental oxygen (15/198 [7.6%] placebo vs 15/399 [3.8%] CAS+IMD **Supplementary Table 6**), consistent with the treatment benefit of CAS+IMD. These events were generally considered by the sponsor as associated with COVID-19 and its complications.

**Table 3.**
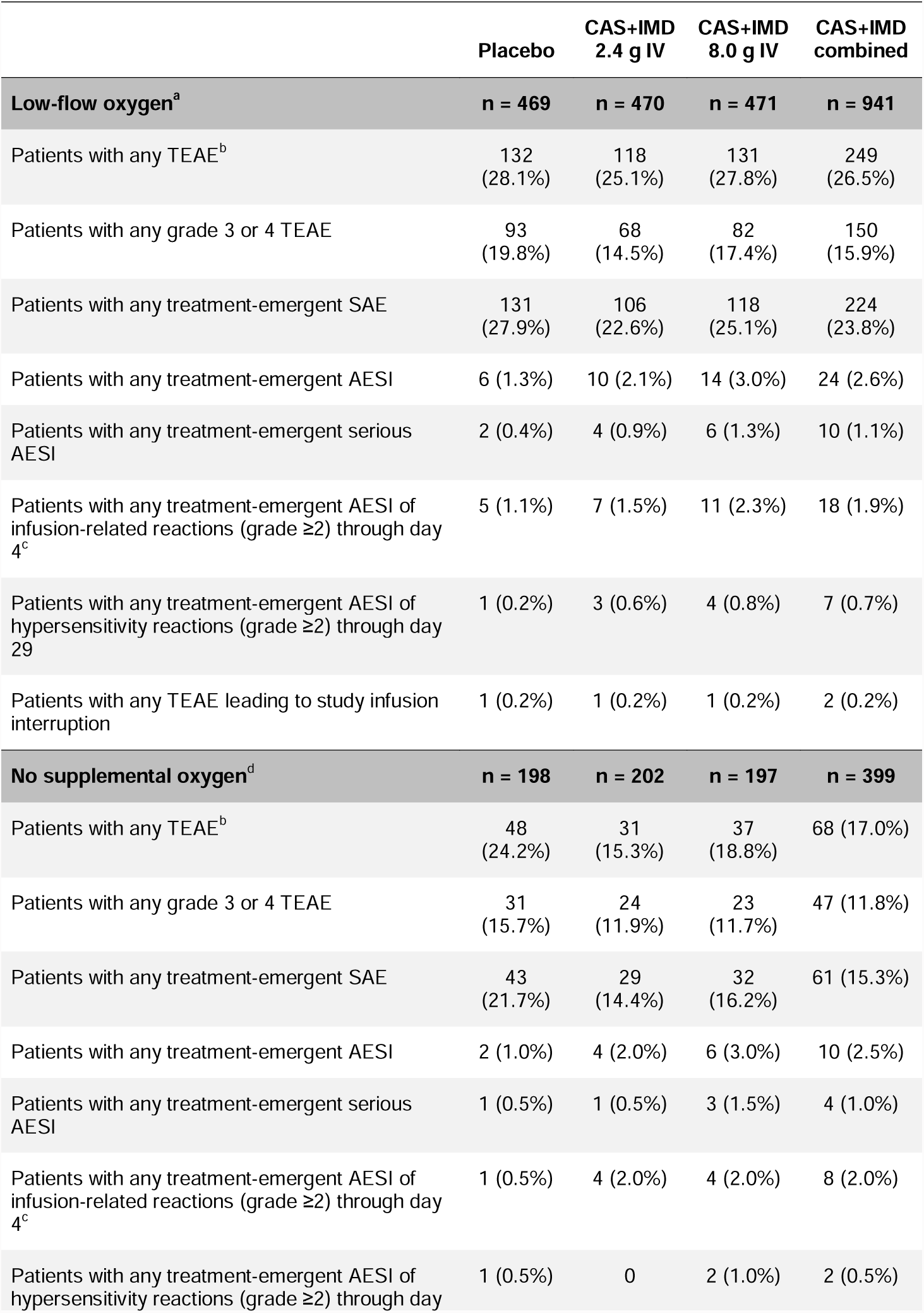
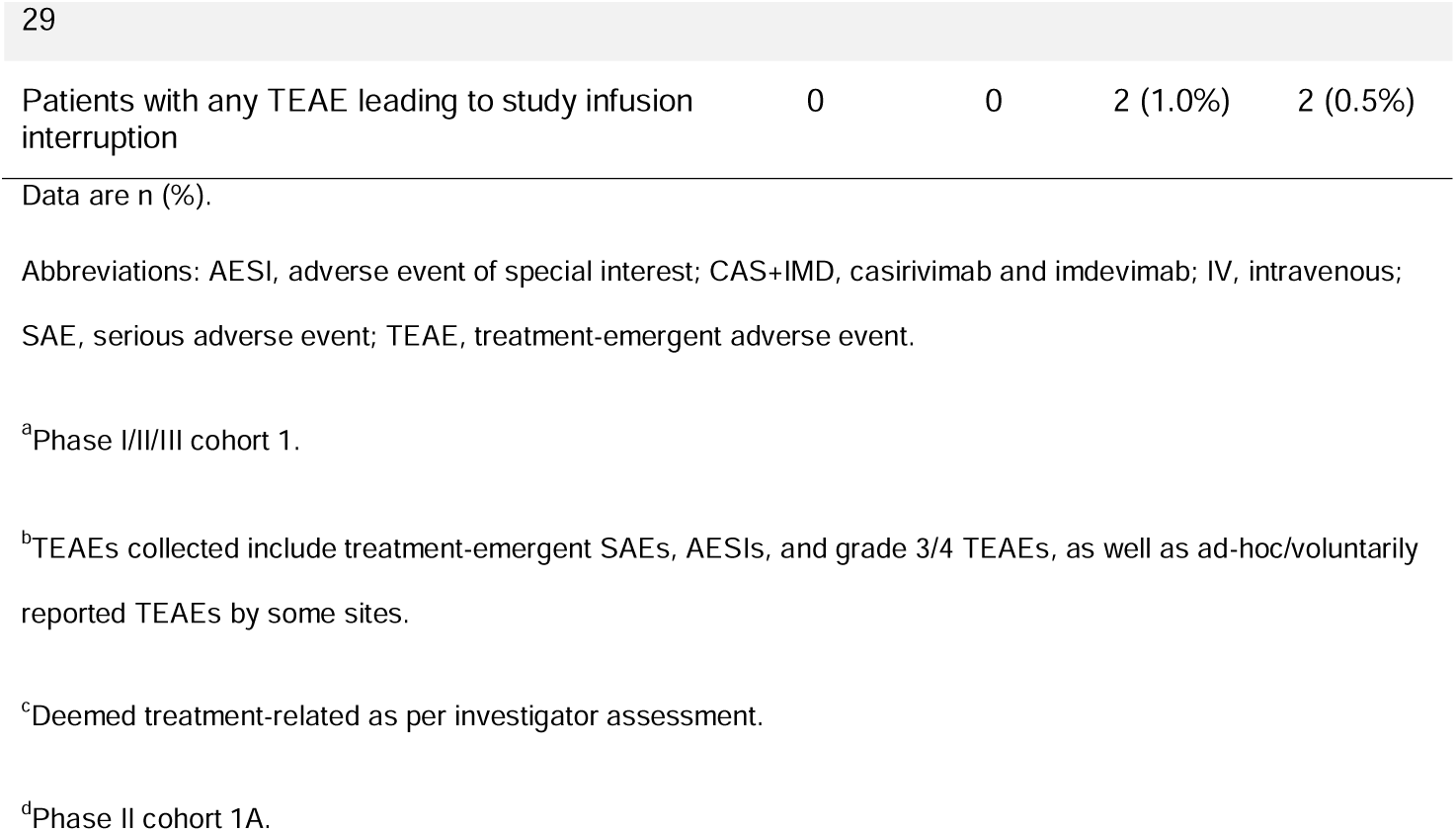
Overview of treatment-emergent adverse events.

Grade ≥2 IRRs occurred in few patients on low-flow (5/469 [1.1%] placebo vs 18/941 [1.9%] CAS+IMD) and no supplemental oxygen (1/198 [0.5%] placebo vs 8/399 [2.0%] CAS+IMD; **Table 3**). Grade ≥2 hypersensitivity reactions also occurred in few patients on low-flow (1/469 [0.2%] placebo vs 7/941 [0.7%] CAS+IMD) and no supplemental oxygen (1/198 [0.5%] placebo vs 2/399 [0.5%] CAS+IMD **Table 3**). AESIs are further detailed in **Supplementary Table 7** and **Supplementary Table 8**.

## Discussion

Hospitalized patients with COVID-19 experience high mortality rates, ranging from 10% to 30% [4, 14-16]. As CAS+IMD is a potent antiviral monoclonal antibody combination shown to rapidly reduce SARS-CoV-2 viral load and modulate a patient’s disease course across various populations [8-11], it was prospectively hypothesized that reducing viral burden as early as possible would decrease morbidity and mortality associated with SARS-CoV-2 hospitalization. Until the recently released results from the RECOVERY platform trial [17], it was unknown whether reducing viral burden in patients who were already hospitalized would be early enough to meaningfully impact clinical outcomes. RECOVERY reported a 20% reduction in 28-day mortality with CAS+IMD treatment compared to standard of care in seronegative patients. The current placebo-controlled randomized trial (with approximately half of patients receiving concomitant remdesivir) demonstrated and extended the benefit preliminarily reported in the RECOVERY trial among seronegative patients, and also documented no harm signals among seropositive patients. When added to standard-of-care treatment, CAS+IMD significantly reduced all-cause mortality for patients on low-flow or no supplemental oxygen. While the primary clinical endpoint, which had a strong positive trend, did not reach significance, all clinical endpoints demonstrated substantial numeric improvements, predominantly driven by the robust and nominally significant results in the seronegative population. In addition, CAS+IMD improved the rates of hospital discharge and death or readmission to hospital at day 29, which persisted through day 57, showing direct benefit to patients as well as the overburdened healthcare system, particularly in the context of viral variants such as delta, which doubles the risk of hospitalization [18].

The clinical benefits observed in the overall population of this study, including improvements in death or mechanical ventilation, all-cause mortality, and discharge from hospital, were driven by seronegative patients, but no harm was demonstrated in seropositive patients, for whom there were numerically fewer deaths through day 29 with CAS+IMD compared to placebo. In the current variant-rich world with widespread COVID-19 vaccination, the utility of serostatus is unclear; numerous publications cite that even vaccinated patients with high antibody titers may have little to no neutralizing activity to emerging variants [19-22]. Given this context, the results from this study may support the use of CAS+IMD in hospitalized patients with COVID-19, regardless of serostatus. Future studies are needed to further explore the potential clinical benefit in seropositive patients.

CAS+IMD is the first monoclonal antibody therapy, and the first SARS-CoV-2 antiviral, that significantly lowers viral load and reduces mortality in hospitalized patients with COVID-19. Very few treatments have demonstrated a mortality benefit in hospitalized COVID-19 patients. Most of these treatments are designed to modulate the immune response late in the disease course after damage has occurred, rather than to clear SARS-CoV-2. The corticosteroid dexamethasone showed a 17% improvement in 28-day mortality in the RECOVERY trial, with the greatest benefit in patients receiving mechanical ventilation [17]. Baricitinib, a Janus kinase inhibitor, improved 28-day mortality by 38% in hospitalized patients [23]. Recently, interleukin-6 inhibitors such as tocilizumab and sarilumab were recommended by the World Health Organization for use in hospitalized patients, in whom they reduced mortality by 13% [24, 25]. The Food and Drug Administration-approved medication remdesivir has shown neither a reduction in viral load nor a mortality benefit in hospitalized patients [26]. CAS+IMD’s mechanism of action and safety profile should allow combination approaches with any or all of these other agents.

The safety profile of CAS+IMD in patients on low-flow or no supplemental oxygen was consistent with that observed previously in outpatients and hospitalized patients with COVID-19 [10, 13], showing low rates of infusion-related and hypersensitivity reactions, as expected for a fully human antibody against an exogenous target. Overall, CAS+IMD was well tolerated. The placebo group experienced a greater frequency of SAEs and adverse events leading to death than the CAS+IMD group, consistent with the clinical benefit of treatment.

The respiratory status of the population in this manuscript includes those receiving low-flow or no supplemental oxygen, as the study did not enroll sufficient numbers of patients on high-intensity oxygen or mechanical ventilation prior to pausing of these cohorts due to an imbalance in mortality observed in interim data early during the conduct of the study, which was not observed in the much larger RECOVERY trial [17]. The absence of full representation across the spectrum of hospitalized patients on varying degrees of oxygen support is a limitation of this study. Additionally, this study was prematurely terminated due to slow recruitment prior to the surge associated with the emergence of the delta variant, and was thus also conducted before the recent B.1.1.529 (omicron) variant began circulating. CAS+IMD, which contains 2 distinct neutralizing antibodies [27, 28], has been shown to have diminished neutralization against the omicron variant [29], but retains neutralizing potency against several viral variants of concern, including B.1.1.7 (alpha), B.1.351 (beta), B.1.617.2 (delta), B.1.429 (epsilon), and P.1 (gamma) [30]. Nonetheless, the results of this trial are informative for circulating variants susceptible to CAS+IMD and could be applicable for the evaluation of future antibodies in the hospitalized population.

As a result of the smaller sample size, key analyses pooled the 2 patient cohorts as well as the 2 doses. Sensitivity analyses did not reveal major efficacy differences across the cohorts or doses; minor variability in the magnitude of risk reductions, with greater effects for the 2.4 g dose compared to the 8.0 g dose, was likely due to small numbers within each group suggesting that either dose can be utilized in hospitalized individuals requiring low-flow or no supplemental oxygen. Overall, trends for treatment benefit on mortality and other efficacy endpoints extend the observations in the larger RECOVERY trial, where benefit was seen in all patients regardless of respiratory status.

Authorized options for intervention with a monoclonal antibody anti-viral treatment have been restricted to outpatients with COVID-19 or as post-exposure prophylaxis [5-7]. Taken together with reports from the RECOVERY trial, these data support CAS+IMD representing a well-tolerated treatment option to reduce the risk of mortality in hospitalized COVID-19 patients and across the disease continuum of SARS-CoV-2, from prevention to hospitalization.

## Supporting information

Cupelli Disclosure Form

Lipsich Disclosure Form

Braunstein Disclosure Form

Stahl Disclosure Form

Bhore Disclosure Form

Hosain Disclosure Form

Ali Disclosure Form

Sivapalasinga Disclosure Form

Somersan Disclosure Form

Pham Disclosure Form

Menon Disclosure Form

Kim Disclosure Form

Soo Disclosure Form

Sun Disclosure Form

Zhao Disclosure Form

Cook Disclosure Form

Mahmood Disclosure Form

DiCioccio Disclosure Form

Hopper Disclosure Form

Kowal Disclosure Form

Pan Disclosure Form

Weinreich Disclosure Form

Mylonakis Disclosure Form

Yancopoulos Disclosure Form

Herman Disclosure Form

Geba Disclosure Form

Wells Disclosure Form

Davis Disclosure Form

Hamilton Disclosure Form

Mei Disclosure Form

Miller Disclosure Form

Turner Disclosure Form

## Data Availability

Qualified researchers may request access to study documents (including the clinical study report, study protocol with any amendments, blank case report form, and statistical analysis plan) that support the methods and findings reported in this manuscript. Individual anonymized participant data will be considered for sharing once the indication has been approved by a regulatory body, if there is legal authority to share the data and there is not a reasonable likelihood of participant re-identification. Submit requests to https://vivli.org/.

## Data sharing

Qualified researchers may request access to study documents (including the clinical study report, study protocol with any amendments, blank case report form, and statistical analysis plan) that support the methods and findings reported in this manuscript. Individual anonymised participant data will be considered for sharing once the indication has been approved by a regulatory body, if there is legal authority to share the data and there is not a reasonable likelihood of participant re-identification. Submit requests to https://vivli.org/.

## Acknowledgements

We thank the patients who participated in this study, as well as their families; the study investigators; the members of the IDMC; Kaitlyn Scacalossi, PhD, and Caryn Trbovic, PhD, from Regeneron Pharmaceuticals for assistance with development of the manuscript; and Prime, Knutsford, UK, for formatting and copyediting suggestions.

