## Supplemental Materials for "Casirivimab and imdevimab for the treatment of hospitalized patients with COVID-19"

**SUPPLEMENTARY APPENDIX**

### Study sites and investigators

**AdventHealth Orlando, Orlando, FL, USA:** Amay Parikh, Jason Sniffen, Wilfred Oniya, Seema Patel

**Ascension Sacred Heart, Pensacola, FL, USA:** Eric Hazbun, Eric Waddington

**Ascension St. Vincent Hospital, Indianapolis, IN, USA:** Francisco Delgado, Markian Bochan, Chad Tewell, Mary Lotz, Hassan Elmalik, Blayre Scott

**Augusta University, Augusta, GA, USA:** Jose Vazquez, Andrew Chao, Budder Siddiqui, Sarah Tran, Caroline Hamilton

**Avera McKennan Hospital and University Health Center, Sioux Falls, South Dakota, USA:** Jawad Nazir, John Lee, Amy Elliott, John Aita

**Banner University Medical Center – Phoenix, Phoenix, AZ, USA:** Marilyn Glassberg, Ramachandra Sista, Raed Alalawi, Esa Rayyan, Thomas Ardiles, Sandra Till, Roland Jabre, Peter Nakaji

**Baylor University Medical Center, Dallas, TX, USA:** Mezgebe Berhe, Sandkovsky Uriel, Dishner Emma, Rahaf Al Masri, Haley Clinton, Erin Duhaime, Christopher Bettacchi

**Boca Raton Regional Hospital, Boca Raton, FL, USA:** Mitchell Karl, Gabriel Sandkovsky, Ashish Neupane, Sanda Cebular, Susan Saxe, Ines Mbaga, Andrew Nguyen, Kurt Wiese

**Boston Medical Center, Boston, MA, USA:** Manish Sagar, Nina Lin, Archana Asundi

**Brigham & Women's Hospital, Boston, MA, USA:** Franscisco Marty, Jennifer Manne-Goehler, Katherine Beluch, Isabel Gonzalez-Bocco, Kaitlyn Timblin

**Carle Foundation Hospital, Urbana, IL, USA:** Karen White, Vishesh Paul, Temitope Shodunke

**Chandler Regional Medical Center, Chandler, AZ, USA:** Chirag Patel, Brian Tiffany

**Clinica Alemana de Santiago, Santiago, Región Metropolitana, Chile:** Sebastián Ibáñez, María Mordojovich, Inia Pérez, Sebastián Solar

**Clínica Las Condes, Las Condes, Santiago de Chile, Chile:** Ricardo Espinoza, Juan Carlos Venegas, Tomás Regueira Heskia, Luisa Durán, Carolina Romero

**Conjunto Hospitalar do Mandaqui, São Paulo, São Paulo, Brazil:** Ana Karolina Barreto Berselli Marinho, Roselene Lourenço

**EME RED Hospitalaria, Mérida, Yucatán, Mexico:** Juan Francisco Rubio Suarez, Ruben Rodrigo Buendia Magaña, Natalia Romero Pavía, Jose Leopoldo Canto Castro, Manuel Alejandro Pasos Mestre, Marco Antonio Aranda Nah

**Emory University, Atlanta, GA, USA:** George Lyon, Delaney Morris, Aneesh Mehta, Colleen Kraft, Nadine Rouphael

**Englewood Health, Englewood, NJ, USA:** Srikant Kondapaneni, Ashwin Jathavedam, Sabena Ramsetty, Aileen Tlamsa, Sejal Gandhi, David Shiu, Ami Vaidya

**Eukarya Pharmasite S.C., Monterrey, Nuevo León, Mexico:** Ricardo Tellez, Sergio Sánchez, Stephani Moreno, Jorge Garza, Alicia López, Alicia Manzo

**Evanston Hospital (NorthShore University Health System), Glenview, IL, USA:** Shashi Bellam, Tom Hensing, Jennifer Grant, Chethra Muthiah

**Hackensack Meridian – Hackensack University Medical Center, Hackensack, NJ, USA:** Steven J. Sperber, Cristina Cicogna,Samit Desai, Lopa Maharaja, Han Nguyen, Anuja Pradhan, Takashi Saito

**Hackensack Meridian – Jersey Shore University Medical Center, Neptune, NJ, USA:** Edward Liu, Jose Fune, Diane Marchesani, Nancy Gornish, Anna Kufelnicka, Takashi Saito

**Harlem Hospital Center – New York City Health and Hospitals Corporation, Harlem, New York, NY, USA:** Farbod Raiszadeh, Sharon Mannheimer, Khaing Myint, Hussein Assallum, Lovelyamma Varghese, Akari Kyaw, Simona Bratu, Raji Ayinla, Anya Weerasinghe

**Holy Name Medical Center, Teaneck, NJ, USA:** Suraj Saggar, Thomas Birch, Benjamin De La Rosa, Karyna Neyra, Erina Kunwar

**Hospital Cardio Pulmonar, Salvador, Bahia, Brazil:** Luiz Eduardo Fonteles Ritt, Marcel Albuquerque, Clarissa Ramos, Ana Tereza Rocha

**Hospital Español Veracruz, Veracruz, Veracruz, Mexico:** Alejandro Quintín Barrat Hernández, Moises Beltrán Molina, Silvano Omar Martínez Pérez, Edgar Iván Muñoz López, Silvia Rios Pérez, Juan Fernando Ramírez Rodriguez, Otto Pinzón Cantanero

**Hospital General de Culiacán “Dr. Bernardo J. Gastélum”, Culiacan, Sinaloa, Mexico:** Jorge Alberto Zamudio Lerma, Elmer Lopez Meza, Minerva Esmeralda Vazquez Huerta, Pablo Andres Reatiga Vega

**Hospital General de Occidente de la Secretaría de Salud Jalisco, Zapopan, Jalisco, Mexico:** Raul Aguilar Orozco, Armando Rafael Vargas Flores, Alicia Elizabeth Guzman Hernandez

**Hospital General Dr. Agustín O´Horan, Mérida, Yucatán, Mexico:** J. Abraham Simón Campos, Felipe de Jesús Pineda Cárdenas, Jessie Carolina Avila Huertas, Jorge Jonatan Angelares Cruz

**Hospital Nossa Senhora das Graças, Porto Alegre, Rio Grande do Sul, Brazil:** Clovis Arns da Cunha, Fernanda Pedroso, Maicon Pinto, Fernanda Staub, Giovanna Brito, Eduardo Malinowski, Denise Hnatiuk, Murilo Guedes, Allan Salva, Guilherme Fioramonte, Gustavo Giavarini, Jessica Souza

**Houston Methodist Hospital, Houston, TX, USA:** Eric Salazar, David Bernard, Brian Castillo, Christopher Leveque, Jian Chen, Cullen Hebert

**Icahn School of Medicine at Mount Sinai, New York, NY, USA:** Judith A. Aberg, Michelle Cespedes, Alexandra Abrams-Downey, Erna Kojic, Luz Lugo, Sean Liu, Nadim Salomon, David Perlman, Deena Altman, Farah Rahman, Georgina Osorio, Joseph Mathew, Sanjana Koshy, Dana Mazo, Francesca Cossarini, Sondra Middleton, Alina Jen, Erika Maria Reategui Schwarz

**Instituto Dante Pazanese de Cardiologia, São Paulo, São Paulo, Brazil:** Diandro Mota, Louis Ohe, David Nunes, Vanessa Salazar, Renata Viana

**Instituto Méderi de Pesquisa e Saúde, Passo Fundo, Rio Grande do Sul, Brazil:** Vinicius Dal Maso, Glaucia Tres, Tiago Simon, Leonardo Priori, Danuza Mello

**Intermountain Medical Center, Murray, UT, USA:** Ithan D. Peltan, Samuel M. Brown, Daanish Hoda, Brandon Webb, Lindsay Leither, Kirk Knowlton

**LDS Hospital, Salt Lake City, UT, USA:** Daanish Hoda, Ithan D. Peltan, Samuel M. Brown, Kirk Knowlton, Lindsay Leither, Brandon Webb

**Jacobi Medical Center, New York, New York, USA:** David Stein, Jason Leider, Gabriele De Vos, Kellie Roe, Jane Devereux, Elizabeth Jenny-Avital, Leonidas Palaiodimos, Ita Nagy Fanny, Julie Hoffman, Jannet Tobon Ramos

**Lincoln Medical Center – New York City Health and Hospitals Corporation, New York, NY, USA:** Vidya Menon, Moiz Kasubhai, Usha Venugopal, Anjana Pillai, Daniel Sittler, Nargis Jilani

**Long Beach Medical Center, Long Beach, CA, USA:** Jimmy Johannes, Thomas Jiang, Christopher Yee, Henry Su, Anthony Arguija

**Maryland School of Medicine, Baltimore, MD, USA:** Uzoamaka Eke, Shivakumar Narayanan, Shyam Kottilil, Joel Chua, Jennifer Husson, John Baddley, R. Gentry Wilkerson

**Mayo Clinic Hospital Rochester, Rochester, MN, USA:** Raymund Razonable, Paschalis Vergidis

**Miami Valley Hospital, Dayton, OH, USA:** Thomas Herchline, Steve Burdette, Jonathan Pope, David Herman

**Morristown Medical Center - AHS Affiliate, Morristown, NJ, USA:** Eric Whitman, Mohamad Cherry, Charles Farber, Jason Kessler, Angela Alistar, Sophie Morse, Robert Roland, Christopher Buck

**Mount Sinai IAM Morningside Clinic, New York, NY, USA:** Judith A. Aberg, Michelle Cespedes, Alexandra Abrams-Downey, Erna Kojic, Luz Lugo, Sean Liu, Nadim Salomon, David Perlman, Deena Altman, Farah Rahman, Georgina Osorio, Joseph Mathew, Sanjana Koshy, Dana Mazo, Francesca Cossarini, Sondra Middleton, Alina Jen, Erika Maria Reategui Schwarz

**Mount Sinai West, New York, NY, USA:** Judith A. Aberg, Michelle Cespedes, Alexandra Abrams-Downey, Erna Kojic, Luz Lugo, Sean Liu, Nadim Salomon, David Perlman, Deena Altman, Farah Rahman, Georgina Osorio, Joseph Mathew, Sanjana Koshy, Dana Mazo, Francesca Cossarini, Sondra Middleton, Alina Jen, Erika Maria Reategui Schwarz

**National Institute for Infectious Diseases “Prof. Dr. Matei Bals”, Bucharest, Romania and Arensia Exploratory Medicine:** Anca Streinu-Cercel, Adrian Streinu, Oana Sandulescu, Ana Maria Andone, Daniela Dospinoiu, Loredana Patru, Magdalena Motoi, Victor Daniel Miron, Alina Alexandra Oana

**Northwest Texas Healthcare System, Amarillo, TX, USA:** David Brabham, Mark Sigler, Manish Patel, Mohammed Al Deen

**Northwestern University, Chicago, IL, USA:** Richard Wunderink, Benjamin Singer, James Walter, Helen Donnelly

**Norton Audubon Hospital, Louisville, KY, USA:** Joseph Flynn, Paul Schulz, Shadi Parsaei, Jeffrey Reeves

**Norton Healthcare, Louisville, KY, USA:** Joseph Flynn, Paul Schulz, Shadi Parsaei, Jeffrey Reeves

**Nuevo Hospital Civil de Guadalajara, Guadalajara, Jalisco, Mexico:** Daniel Rodriguez Gonzalez, Ileana Higareda Almaraz, Victor Hugo Madrigal

**OhioHealth Riverside Methodist Hospital, Columbus, OH, USA:** Brian Zeno, Jessica Kynyk, Edward Cordaso, Zachary Baird

**Overlook Medical Center, Summit, NJ, USA:** Eric Whitman, Sophie Morse, Robert Roland

**Parkland Hospital and Health System, Dallas, TX, USA:** Trushil Shah, Catherine Chen, Reuben Arasaratnam

**PharmaTex Research, LLC, Amarillo, TX, USA:** David Brabham, Mark Sigler, Tarek Naguib

**Piedmont Healthcare, Atlanta, GA, USA:** Amy Case, Chad Miller, Craig Patterson, Raymond Rubin, Christine Zurawski

**Providence - Covenant Hospital, Lubbock, TX, USA:** Dennis Duriex, Prakash Shrestha

**Providence Holy Cross Medical Center, Mission Hills, CA, USA:** Aric Gregson, Kasra Sedarati, Alan Nazarian, Sasan Sani, Babak Eshaghian

**Providence Portland Medical Center, Portland, OR, USA:** Jason Wells, David Hotchkin, Jeffery Robinson, Hataya Poonyagariyagorn, Tobias Pusch, Jennifer Marfori, Cameron Cover, Brian Kendall, Nicholas Stucky, Amy Dechet, Justin Jin

**Providence Regional Medical Center Everett, Everett, WA, USA:** George Diaz, Daniel McClung, Robert Choi, Albert Pacifico

**Providence Saint John's Health Center, Santa Monica, CA, USA:** Terese Hammond, Fabian Andres Romero, Steven O'Day, Trevan Fischer, Ana Rocha, Anmol Rangoola

**New York City Health + Hospitals/Queens, Jamaica, NY, USA:** Jazila Mantis, Margaret Kemeny, Merjona Saliaj

**Raymond G. Murphy Veterans Affairs Medical Center, Albuquerque, NM, USA:** Sarah Medrek, Karen Servilla

**PMSI Republican Clinical Hospital “T. Mosneaga”, ARENSIA EM, Chișinău, Moldova:** Victor Cojocaru, Alexandru Botizatu, Dinu Condrea, Daniel Mindrila, Nelea Ghicavii, Angela Coltuclu, Sergiu Ursul, Arteom Zarisneac

**Rhode Island Hospital, Providence, RI, USA:** Eleftherios Mylonakis, Ralph Rogers, Fadi Shehadeh, Saisanjana Kalagara, Maria Tsikala-Vafea, Eleftheria Atalla, Evangelia K. Mylona, Matthew Kaczynski, Biswajit Mishra, Lewis Felix Raj Lucas

**Rush University Medical Center, Chicago, IL, USA:** Graeme Forrest, Mariam Aziz, Carlos Santos, Shivanjali Shankaran, Nipun Atri

**Sarasota Memorial Hospital, Sarasota, FL, USA:** Manuel Gordillo, Rishi Bhattacharyya, Sudha Tallapragada, Annette Artau, Julie Larkin, Roberto Mercado, Michael Milam, Natan Kraitman, Rabih Loutfi, Kirk Voelker, Lenka Offner, Michael Lowry

**South Shore Infectious Disease and Travel Medicine Consultants & Antibiotic Infusion Center, West Islip, NY, USA:** Uzma Syed, Michael Gray

**Spectrum Health, Grand Rapids, MI, USA:** Gordana Simeunovic, Nicholas Hartog, Derek Vanderhorst, Angela Peraino, Jacob Baker, James Polega, Patricia Choi, Josh Donkin

**Stanford University (Stanford Health), Stanford, CA, USA:** Aruna Subramanian, Philip Grant, Anne Liu, Angela Rogers, John Kugler, Shanthi Kappagoda

**St. Luke's Hospital – Missouri, Chesterfield, MO, USA:** Neil Ettinger, Bobby Shah, Kristen Fisher

**SUNY Upstate Medical University, Syracuse, NY, USA:** Kristopher Paolino, Elizabeth Harausz, Stephen Thomas, Elizabeth Asiago-Reddy, Timothy Endy, Rachael Cavelli

**Supera Oncologia, Chapecó, Santa Catarina, Brazil:** Cristiano Devenci Vendrame, Hugo Silva, Carine Kolling, Aland Waldow, Gabriel Alessio, Paulo Fachinello, Raulério Papini

**Swedish Medical Center, Seattle, WA, USA:** Jason D. Goldman, William Berrington, Mary Micikas, Gregory Moss, Reda Tipton, Allison Everett, Julie Wallick

**Temple University Hospital, Philadelphia, PA, USA:** Gerard Criner, Nathaniel Marchetti, Parag Desai, Daniel Salerno, Fredric Jaffe, Samuel Krachman, Matthew Zheng, Maulin Patel, Junad Chowdhury, Daniel Mueller, Parth Rali, Eduardo Dominguez Castillo, Zachariah Dorey-Stein, John Scott

**Therapeutic Concepts, PA, Houston, TX, USA:** Joseph Gathe Jr., Carl Mayberry, Joseph Varon

**The Ohio State University Wexner Medical Center, Columbus, OH, USA:** Matthew C. Exline, Mohammad Mahdee E. Sobhanie, Joshua A. Englert, Sonal R. Pannu

**The Miriam Hospital, Providence, RI, USA:** Eleftherios Mylonakis, Ralph Rogers, Fadi Shehadeh, Saisanjana Kalagara, Maria Tsikala-Vafea, Eleftheria Atalla, Evangelia K. Mylona, Matthew Kaczynski, Biswajit Mishra, Lewis Felix Raj Lucas

**University at Buffalo, Buffalo, NY, USA:** John K. Crane, Jonathan Claus, Manoj Mammen

**The University of Texas Health Science Center, Tyler, TX, USA:** Julie Philley, Megan Devine

**Tufts Medical Center, Boston, MA, USA:** Brian Chow, Debra Poutsiaka, Roberto Viau Colindres, Andrew Strand, Jenn Chow, Jose Caro, Rakhi Kohli, Malla Bipin, Saba Mostafavi, Christhian Alejandro Cano Guerra, Paula Dabenigno, Vidya Iyer, Yoav Golan, Tine Vindenes, Laura Kogelman, Whitney Perry, Carlos Mendoza

**Tulane University School of Medicine, New Orleans, LA, USA:** Dahlene Fusco, Arnaud Drouin

**Universidade Estadual Paulista, Botucatu, São Paulo, Brazil:** Marina Politi Okoshi, Carolina Rodrigues Tonon, Bertha Furlan Polegato, Luana Urbano Pagan

**University of Alabama at Birmingham, Birmingham, AL, USA:** Sonya Heath, Donna Cote, Paul Goepfert, Tai' Turner-Green, Turner Overton

**University of Arizona, Tucson, AZ, USA:** Sachin Chaudhary, Sairam Parthasarathy

**University of California Davis, Sacramento, CA, USA:** Timothy Albertson, Nicholas Kenyon, Christian Sandrock, Stuart Cohen, Brian Morrisey

**University of Colorado, Aurora, CO, USA:** Thomas Campbell, Amiran Baduashvili, Esther Benamu, Hilary Dunlevy, Suzanne Fiorillo, Steven Johnson, Martin Krsak, Poornima Ramanan

**University of Florida, Gainesville, FL, USA:** Mark Brantly, Ali Ataya, Lisa Merck, Kiran Lukose, Juan Kattan, Kartikeya Cherabuddi, Erin Silverman

**University of Iowa, Iowa City, IA, USA:** Alejandro Comellas, Joel Kline, Spyridon Fortis

**University of Nebraska Medical Center, Omaha, NE, USA:** Diana Florescu, Erica Stohs, Andrea Zimmer, Adia Sikyta, Elizabeth Schnaubelt

**University of North Carolina at Chapel Hill, Chapel Hill, NC, USA:** William Fischer, David Wohl, Anne Lachiewicz, David Margolis, Joseph Eron, Brian Bramson, Nikolaos Mavrigiorgos

**University of Rochester Medical Center, Rochester, NY, USA:** Christopher Palma, Lisa Beck, Ummara Shah

**University of São Paulo, São Paulo, São Paulo, Brazil:** Esper G. Kallas, Angela Freitas, Jessica F. Ramos

**University of South Florida, Tampa, FL, USA:** Kami Kim, Seetha Lakshmi, Tiffany Vasey, Lucy Guerra, Susannah Hall

**University of Texas Southwestern Medical Center, Dallas, TX, USA:** Trushil Shah, Mamta Jain, Corey Kershaw, Leah Cohen, Catherine Chen, David Finklea, Kelly Chin, Peiman Lahsaei, Edward Mims

**University of Wisconsin, Madison, WI, USA:** William Hartman, Joseph Connor, Robert Striker, Kraig Kumfer, Nicole Bonk, Shahzeb Munir, David Sterkin, Scott Ensminger, Jashan Octain, Ann Sheehy, Alexis Waters, Scott Wilson

**University Medical Center New Orleans (LCMC Health), New Orleans, LA, USA:** Dahlene Fusco, Arnaud Drouin, Joshua Denson, Jerry Zifodya, Christine Bojanowski, Celeste Newby

**VA Western New York Healthcare System at Buffalo, Buffalo, NY, USA:** Karin Provost, Archana Mishra, M. Jeffery Mador, Gregory Fuhrer

**VA Portland Healthcare System, Portland, OR, USA:** Mitchell Sally, Graeme Forrest, Matthew Diveronica, Thomas Barrett

**Virginia Commonwealth University School of Medicine, Richmond, VA, USA:** Marjolein de Wit, Patrick Nana-Sinkam, Aamer Syed

**Washington University in St Louis, St. Louis, MO, USA:** Andrej Spec, Rachel Presti, Jane O'Halloran, Carlo Mejia, Patrick Mazi, Adriana Rauseo Acevedo

**Wellstar Kennestone Hospital, Marietta, GA, USA:** Danny Branstetter, Neha Paranjape

**White Plains Hospital Center, White Plains, NY, USA:** Jennifer Schelker, Peter Chu, Shyam Vadlapatla, Dan Sammartino, Jessica Maldonado, Mirjam Norris-Nommensen, Kimberly Farrell, Cigi Mathew

### Regeneron study team

A. Thomas DiCioccio, Adebiyi Adepoju, Adnan Mahmood, Aisha Mortagy, Ajla Dupljak, Alison Brown, Alpana Waldron, Amanda Cook, Amra Arslanagic, Amy Froment, Andrea T. Hooper, Andrea Margiotta, Anita Islam, Anne Smith, Arvinder Dhillon, Aurora Breazna, Bari Kowal, Barry Silverstein, Bret Musser, Brian Bush, Brian Head, Bryan Zhu, Camille Debray, Careta Phillips, Carmella Simiele, Carol Lee, Carolyn Nienstedt, Caryn Trbovic, Catherine Elliott, Chad Fish, Charlie Ni, Charlotte Lyon, Christa Polidori, Christina Perry, Christine Enciso, Christopher Chamak, Christopher Powell, Cynthia Pan, Dana Wolken, Danise Subramaniam, David Liu, David M. Weinreich, David Stein, Dawlat Hassan, Daya Gulabani, Deborah Fix, Deborah Leonard, Deepshree Sarda, Denise Bonhomme, Denise Kennedy, Derrick Bramble, Devin Darcy, Dhanalakshmi Barron, Diana Hughes, Diana Rofail, Dipinder Kaur, Dominique Atmodjo Watkins, Dona Bianco, Donna Gambaccini, Eduardo Forleo Neto, Edward Jean-Baptiste, Ehsan Bukhari, Elizabeth Bucknam, Emily Nanna, Esther Huffman O'Keefe, Evelyn Gasparino, Evonne Fung, Flonza Isa, Fung-Yee To, Gary Herman, Gayatri Anand, George D. Yancopoulos, Georgia Bellingham, Giane Sumner, Grainne Moggan, Grainne Power, Gregory P. Geba, Gwyn Dixon, Haixia Zeng, Heath Gonzalez, Helen Cicirello, Helen Kang, Hibo Noor, Ian Minns, James Donohue, Jamie Rusconi, Janice Austin, Janie Parrino, Jeannie Yo, Jenna McDonnell, Jennifer D. Hamilton, Jessica Boarder, Jianguo Wei, Jing Xiao, Jingchun Yu, Jingning Mei, Joanne Malia, Joanne Tucciarone, Jodie Tyler-Gale, John D. Davis, John Rembis, John Strein, Jonathan Cohen, Jonathan Meyer, Jordan Ursino, Joseph Im, Joseph Tramaglini, Joseph Wolken, Jutta Miller, Kaitlyn Potter, Kaitlyn Scacalossi, Kamala Naidu, Kara Ford, Karen Browning, Karen Yau, Katherine Woloshin, Kelly Lewis-Amezcua, Kenneth C. Turner, Kit Chiu, Kristina McGuire, Kristy Macci, Kurt Ringleben, Kyle Foster, Lacey Douthat, Latora Knighton, Leah Lipsich,^†^ Lillian Brener, Linda Kelly, Lindsay Darling, Lisa Boersma, Lisa Cowen, Lisa Cupelli, Lisa Hersh, Lisa Jackson, Lisa Purcell, Lisa Sherpinsky, Lori Geissler, Louise Boppert, Lyra Fiske, Mahesh Vadyala, Manika Bista, Marc Dickens, Maureen Weimer, Meagan O'Brien, Michael Batchelder, Michael Partridge, Michel Tarabocchia, Mivia Rodriguez, Moetaz Albizem, Muriel O'Byrne, Nagaratna Medapati, Ned Braunstein, Neena Sarkar, Neil Stahl, Ngan Trinh, Nicholas Moore, Nicole Deitz, Nicole Memblatt, Nirav Shah, Nitin Kumar, Nkechi Moghalu, Olga Herrera, Oluchi Adedoyin, Ori Yellin, Pamela Snodgrass, Patrick Floody, Paul D'Ambrosio, Peter Boutros, Prankur Krishnatry, Qin Li, Rafia Bhore, Rakiyya Ali, Ramya Iyer, Rinol Alaj, Rita Pedraza, Robert Hamlin, Romana Hosain,^†^ Ruchin Gorawala, Ryan White, Ryan Yu, Rylee Fogarty, S. Balachandra Dass, Sagarika Bollini, Samit Ganguly, Sandra DeCicco, Sandra Osbild, Sara Dale, Selin Somersan-Karakaya, Sharon Henkel, Shazia Ali, Shelley Geila Shapiro, Soraya Nossoughi, Steve Chen, Steven Elkin, Steven Long, Sumathi Sivapalasingam,^†^ Susan Irvin, Susan Wilt, Suzanne Luther, Tami Min, Tatiana Constant, Theresa Devins, Travis Bernardo, Viet Pham, Violet Vincent, Xin Chen, Yanmei Tian, Yasmin Khan, Yiping Sun, Yuhwen Soo, Yuming Zhao, Yunji Kim

^†^Former employee of Regeneron Pharmaceuticals, Inc.

### Supplementary methods

#### Inclusion and exclusion criteria

##### Inclusion criteria

A patient must meet the following criteria to be eligible for inclusion in the study:

1. Has provided informed consent (signed by study patient or legally acceptable representative)
2. Male or female adult ≥18 years of age (or country’s legal age of adulthood if higher) at randomization
3. Has severe acute respiratory syndrome coronavirus 2 (SARS-CoV-2)-positive antigen or molecular diagnostic test (by validated SARS‑CoV-2 antigen, quantitative reverse transcription polymerase chain reaction [RT-qPCR], or other molecular diagnostic assay, using an appropriate sample such as nasopharyngeal [NP], nasal, oropharyngeal, or saliva) ≤72 hours prior to randomization and no alternative explanation for current clinical condition. A historical record of positive result from test conducted ≤72 hours prior to randomization is acceptable.
4. Has symptoms consistent with coronavirus disease 2019 (COVID-19), as determined by investigator, with onset ≤10 days before randomization
5. Hospitalized for ≤72 hours with at least 1 of the following at randomization; patients meeting more than 1 criterion will be categorized in the most severely affected category:
   a. **Cohort 1A (no supplemental oxygen):** With COVID-19 symptoms but not requiring supplemental oxygen
   b. **Cohort 1 (low-flow oxygen):** Maintains O_2_ saturation >93% on low-flow oxygen via nasal cannula, simple face mask, or other similar device

*Note: Sites located in high-altitude areas (>1500 m above sea level) should refer to appendix A of the Clinical Study Protocol for the appropriate high-altitude equivalents for sea-level oxygenation measurements.*

c. **Cohort 2^a^ (high-intensity oxygen):** High-intensity oxygen therapy without mechanical ventilation, where high-intensity is defined as receiving supplemental oxygen delivered by one of the following devices:
- Non-rebreather mask (with SpO_2_ ≤96% while receiving an oxygen flow rate of at least 10 L/min)
- High-flow device (eg, AIRVO^™^ or Optiflow^™^) with at least 50% FiO_2_
- Non-invasive ventilator, including continuous positive airway pressure to treat hypoxemia (excluding isolated use for sleep-disordered breathing)
d. **Cohort 3^a^ (mechanical ventilation):** On mechanical ventilation

*^a^Per independent data monitoring committee (IDMC) recommendation first received on October 30, 2020, and reiterated through February 19, 2021, patient enrolment in cohort 2 and cohort 3 was placed on hold.*

##### Exclusion criteria

A patient who meets any of the following criteria will be excluded from the study:

1. **Phase I only:** Patients maintaining O_2_ saturation >94% on room air

*Note: For sites in high-altitude areas, refer to appendix A of the Clinical Study Protocol. Patients on room air will not be excluded from phase II.*

2. In the opinion of the investigator, unlikely to survive for >48 hours from screening
3. Receiving extracorporeal membrane oxygenation
4. Has new-onset stroke or seizure disorder during hospitalization
5. Initiated on renal replacement therapy due to COVID-19
6. Has circulatory shock requiring vasopressors at randomization

*Note: Patients who require vasopressors for sedation-related hypotension or reasons other than circulatory shock may be eligible in this study.*

7. Patients who have received convalescent plasma, intravenous immunoglobulin (IVIG), or monoclonal antibodies against SARS-CoV-2 (eg, bamlanivimab) within 5 months prior to randomization or plan to receive during the study period for any indication
8. Participation in a clinical research study, including any double-blind study, evaluating an investigational product within 30 days and fewer than 5 half-lives of the investigational product prior to the screening visit

*Note: The use of remdesivir, hydroxychloroquine, or other treatments (except for COVID-19 convalescent plasma or IVIG) being used for COVID-19 treatments in the context of the local standard of care or an open-label study or compassionate use protocol is permitted.*

9. Any physical examination findings, history of illness, and/or concomitant medications that, in the opinion of the study investigator, might confound the results of the study or pose an additional risk to the patient by their participation in the study
10. Known allergy or hypersensitivity to components of study drug
11. Pregnant or breastfeeding women
12. Continued sexual activity in women of childbearing potential (WOCBP)^a^ or sexually active men who are unwilling to practice highly effective contraception prior to the initial dose/start of the first treatment, during the study, and for at least 6 months after the last dose

Highly effective contraceptive measures in WOCBP include:

- Stable use of combined (estrogen- and progestogen-containing) hormonal contraception (oral, intravaginal, transdermal) or progestogen-only hormonal contraception (oral, injectable, implantable) associated with inhibition of ovulation initiated 2 or more menstrual cycles prior to screening
- Intrauterine device
- Intrauterine hormone-releasing system
- Bilateral tubal ligation
- Vasectomised partner,^b^ and/or
- Sexual abstinence^c,d^

Male study participants with WOCBP partners are required to use condoms unless they are vasectomized^b^ or practice sexual abstinence.^c,d^

^a^WOCBP are defined as women who are fertile following menarche until becoming postmenopausal, unless permanently sterile. A postmenopausal state is defined as no menses for 12 months without an alternative medical cause. A high follicle-stimulating hormone (FSH) level in the postmenopausal range may be used to confirm a postmenopausal state in women not using hormonal contraception or hormonal replacement therapy. However, in the absence of 12 months of amenorrhea, a single FSH measurement is insufficient to determine the occurrence of a postmenopausal state. The above definitions are according to Clinical Trial Facilitation Group guidance. Pregnancy testing and contraception are not required for women with documented hysterectomy or tubal ligation. Permanent sterilization methods include hysterectomy, bilateral salpingectomy, and bilateral oophorectomy.

^b^Vasectomized partner or vasectomized study participant must have received medical assessment of the surgical success.

^c^Sexual abstinence is considered a highly effective method only if defined as refraining from heterosexual intercourse during the entire period of risk associated with the study drugs. The reliability of sexual abstinence needs to be evaluated in relation to the duration of the clinical trial and the preferred and usual lifestyle of the patient.

^d^Periodic abstinence (calendar, symptothermal, post-ovulation methods), withdrawal (coitus interruptus), spermicides only, and lactational amenorrhea method are not acceptable methods of contraception. Female condom and male condom should not be used together.

#### SARS-CoV-2 serostatus determination

All patients were assessed for baseline viral load and the presence or absence of anti-SARS-CoV-2 antibodies: anti-spike (S1) immunoglobulin (Ig) A, anti-S1 IgG, and anti-nucleocapsid IgG. Patients underwent randomization regardless of their baseline serostatus, and were grouped for analyses as seronegative (if all baseline antibody tests were negative), seropositive (if any baseline antibody test was positive), borderline (if any test was borderline), or other (missing, not determined, pending, or inconclusive results). For analysis, “borderline” cases were categorized as “other.”

#### Patient cohorts

At randomization, patients were enrolled in 1 of f4our cohorts based on disease severity. Patients meeting more than 1 criterion were categorized in the most severely affected category:

- **Cohort 1A (no supplemental oxygen)**: Patients with COVID-19 symptoms but not requiring supplemental oxygen
- **Cohort 1 (low-flow oxygen)**: Patients with O_2_ saturation >93% on low-flow oxygen via nasal cannula, simple face mask, or other similar device
- **Cohort 2 (high-intensity oxygen)**: Patients on high-intensity oxygen therapy but not on mechanical ventilation
  - High-intensity oxygen therapy is defined as the use of non-rebreather mask with an oxygen flow rate of at least 10 L/min, use of a high-flow device with at least 50% FiO_2_, or use of non-invasive ventilation to treat hypoxemia
- **Cohort 3 (mechanical ventilation)**: Patients on mechanical ventilation

This manuscript presents data from cohort 1A and cohort 1. Enrolment into cohort 2 and cohort 3 was paused per IDMC recommendation, as described in the trial adaptions section of the appendix, and was not resumed prior to termination of the study by the sponsor.

#### Trial adaptations

The phase I portion of the study enrolled patients receiving low-flow oxygen only. Initiation of the phase II portion of the trial was contingent upon IDMC review of phase I data from the sentinel safety group, after which patients were enrolled into all 4 cohorts.

On October 30, 2020, the IDMC recommended pausing enrolment of patients receiving high-intensity oxygen (cohort 2, n = 161) and mechanical ventilation (cohort 3, n = 35) based on an imbalance in mortality observed in interim data early during the conduct of the study (**Supplementary Table 9**, **Supplementary Table 10**). Enrolled patients in the discontinued cohorts completed the study follow-up but, due to the very limited numbers in these cohorts, these data are not included in the current analysis. This pause was maintained for the remainder of the study, while the IDMC recommended proceeding with enrolment of patients receiving low-flow (cohort 1) or no (cohort 1A) supplemental oxygen.

On April 9, 2021, a decision was made by the sponsor to terminate further patient enrolment in this study, which at the time was still enrolling cohorts 1 and 1A while cohorts 2 and 3 were paused. This decision was made for strategic reasons and was not based on any safety concerns: enrolment was proceeding very slowly (prior to the surge associated with the delta variant), and the much larger RECOVERY trial had just announced a mortality benefit.[1] Until this time, only patients receiving low-flow oxygen (cohort 1) were enrolled into the phase III portion of the study. Accordingly, enrolment of patients receiving low-flow (cohort 1, phase III) and no (cohort 1A, phase II) supplemental oxygen was prematurely terminated. All ongoing patients were followed up through the end of the study.

#### Trial oversight

Regeneron Pharmaceuticals, Inc. designed the trial and, with the trial investigators, gathered the data. Regeneron Pharmaceuticals, Inc. analyzed the data. The summary of protocol amendments is available in the protocol and a complete list of trial investigators is presented in the appendix. The investigators, site personnel, and Regeneron Pharmaceuticals, Inc. were unaware of the treatment-group assignments. An IDMC monitored unblinded data to make recommendations about trial modifications.

The trial was conducted in accordance with the principles of the Declaration of Helsinki, International Council for Harmonisation Good Clinical Practice guidelines, and applicable regulatory requirements. The local institutional review board or ethics committee at each study center oversaw trial conduct and documentation. All patients provided written informed consent before participating in the trial.

#### Additional statistical methods

##### Missing data handling

The virologic endpoint analysis was based on the observed data with no imputation for missing data. For the time-weighted average (TWA) change from baseline calculation in the primary virologic endpoint, if the change from baseline in viral load was missing due to failed test or other reasons, only the time points with non-missing values were included in the calculation. Patients with a missing baseline value were excluded from the analysis. In the analysis for the clinical endpoints based on proportions of patients with the event, the missing data will be considered as non-events.

##### RT-qPCR parameters

The validated RT-qPCR assay for SARS-CoV-2 utilized was previously described (Viracor-Eurofins, USA).[2] The limit of detection was 714 copies/mL.

##### Sample size calculations

The phase II sample size was estimated to achieve 80% power to detect a risk reduction of 50% (hazard ratio=0.5) between any CAS+IMD dose group (2.4 g or 8.0 g) versus placebo at α=0.1 (1-sided). The phase III sample size was calculated assuming that 13.1% of placebo patients died or went on mechanical ventilation from days 1 to 29 and α=0.05 (2-sided); therefore, the minimal significant differences in relative risk reduction between the CA+IMD combined dose group and placebo group were 29.0%, 41.2%, and 36.6% for the overall modified full analysis set (mFAS), seronegative mFAS, and high-viral load mFAS patients, respectively. Sample sizes were not recalculated following premature termination of the study.

##### Primary virologic endpoint analysis

The primary virologic efficacy endpoint of TWA change from baseline viral load in NP samples through day 7 was performed in seronegative patients (seronegative mFAS) not requiring supplemental oxygen or on low-flow oxygen. Virologic efficacy was analyzed using the analysis of covariance model with treatment group and the type of background standard of care as fixed effects, and baseline viral load and treatment by baseline interaction as covariates, as predefined in the statistical analysis plan.

##### Primary clinical endpoint analysis

The primary clinical efficacy endpoint of death or mechanical ventilation was estimated by the proportion of patients who died or went on mechanical ventilation from day 6 through day 29 and from day 1 through day 29 for patients not requiring supplemental oxygen or on low-flow oxygen in the following subgroups: high-viral load (baseline viral load >10^6^ copies/mL) mFAS, seronegative mFAS, and mFAS (also referred to as the high-viral load population, seronegative population, and overall population). Clinical efficacy was analyzed using the landmark analysis approach for day 6 through day 29, as well as for day 1 through day 29. The proportion of patients who died or went on mechanical ventilation was analyzed using either the exact method for binomial distribution or asymptotic normal approximation method, based on the number of events. If the number of events was small (eg, np ≤5 or n(1-p) ≤5 in any treatment group, where n is the number of patients in the treatment group and p is the proportion of events), then the Fisher’s exact test was applied. Otherwise the Cochran–Mantel–Haenszel test, stratified by the type of background standard of care (antiviral therapies and non-antiviral therapies), was applied. Relative risk reduction (RRR; 1 minus relative risk) and corresponding 95% confidence intervals compared to the placebo group were estimated by the Farrington-Manning method.

### Supplementary results

##### Discharge from hospital

CAS+IMD led to numeric improvements in the proportion of seronegative patients discharged alive from the hospital: 324/360 patients (90.0%) were discharged in the CAS+IMD group, compared to 130/160 patients (81.2%) in the placebo group (RRR, –10.8% to 95% confidence interval [CI]: –20.2% to –2.0%; nominal *P* = 0.0072 **Figure 2B**). Consistent results were observed for hospital discharge in the overall population (714/804 [88.8%] CAS+IMD vs 330/393 [84.0%] placebo; RRR, –5.8%; 95% CI: –11.1% to –0.6%; nominal *P* = 0.0184; **Figure 2B**). The median time to discharge from hospital was 4 days (95% CI: 4.0–5.0) for patients in both the placebo and CAS+IMD group, whereas the 75% quantile time to discharge was 11 days (95% CI: 10.0–15.0) for placebo versus 9 days (95% CI: 8.0–10.0) for CAS+IMD (**Supplementary Table 3**).

##### Death or readmission to hospital

CAS+IMD led to numeric reductions in death or readmission to hospital at day 29 in seronegative patients (44/360 [12.2%] CAS+IMD vs 39/160 [24.4%] placebo; RRR, 49.9%; 95% CI: 26.0–66.0%). This improvement persisted thorough day 57 (65/360 [18.1%] CAS+IMD vs 46/160 [28.8%] placebo; RRR, 37.2%; 95% CI: 12.8–54.8%; **Supplementary Table 4**). The relative risk reductions in the overall population (RRR, 24.9%; 95% CI: 0.3–43.4%) and high-viral load population (RRR, 22.4%; 95% CI: –7.4% to 43.9%) at day 29 showed a similar trend with lower magnitude (**Supplementary Table 4**).

Supplementary Table 1. Statistical hierarchy

| Type | Description | Testing order |
| --- | --- | --- |
| Primary virologic outcome | Time-weighted average change from baseline viral load in NP sample through day 7 **in the seronegative** **mFAS** for comparing the combined doses of CAS+IMD versus placebo | 1 |
| Primary clinical outcome | Proportion of patients who died or went on mechanical ventilation **from day 6 to day 29 in the high-viral load mFAS** for comparing the combined doses of CAS+IMD versus placebo | 2 |
|  | Proportion of patients who died or went on mechanical ventilation **from day 6 to day 29 in the seronegative** **mFAS** for comparing the combined doses of CAS+IMD versus placebo | 3 |
|  | Proportion of patients who died or went on mechanical ventilation **from day 6 to day 29 in the overall mFAS** for comparing the combined doses of CAS+IMD versus placebo | 4 |
|  | Proportion of patients who died or went on mechanical ventilation **from day 1 to day 29 in the high-viral load mFAS** for comparing the combined CAS+IMD versus placebo | 5 |
|  | Proportion of patients who died or went on mechanical ventilation **from day 1 to day 29 in the seronegative mFAS** for comparing the combined doses of CAS+IMD versus placebo | 6 |
|  | Proportion of patients who died or went on mechanical ventilation **from day 1 to day 29 in the overall mFAS** for comparing the combined doses of CAS+IMD versus placebo | 7 |

Abbreviations: CAS+IMD, casirivimab and imdevimab; mFAS, modified full analysis set; NP, nasopharyngeal.

Supplementary Table 2. Demographics and baseline characteristics by baseline serum antibody status for patients on low-flow or no supplemental oxygen

|  | **Placebo** | **CAS+IMD  2.4 g IV** | **CAS+IMD  8.0 g IV** | **CAS+IMD  combined doses** | **Total** |
| --- | --- | --- | --- | --- | --- |
| **Seronegative^a^** | **n = 160** | **n = 172** | **n = 188** | **n = 360** | **n = 520** |
| Age |  |  |  |  |  |
| Median (range) | 64.0 (26–96) | 60.0 (23–97) | 63.0 (26–98) | 62.0 (23–98) | 62.5 (23–98) |
| ≥65 | 76 (47.5%) | 75 (43.6%) | 87 (46.3%) | 162 (45.0%) | 238 (45.8%) |
| Male sex | 82 (51.3) | 96 (55.8) | 101 (53.7) | 197 (54.7) | 279 (53.7) |
| Race |  |  |  |  |  |
| White | 93 (58.1%) | 107 (62.2%) | 132 (70.2%) | 239 (66.4%) | 332 (63.8%) |
| Black or African American | 18 (11.3%) | 21 (12.2%) | 27 (14.4%) | 48 (13.3%) | 66 (12.7%) |
| Asian | 7 (4.4%) | 6 (3.5%) | 2 (1.1%) | 8 (2.2%) | 15 (2.9%) |
| American Indian or Alaska Native | 1 (0.6%) | 2 (1.2%) | 2 (1.1%) | 4 (1.1%) | 5 (1.0%) |
| Native Hawaiian or Pacific Islander | 0 | 1 (0.6%) | 1 (0.5%) | 2 (0.6%) | 2 (0.4%) |
| Unknown | 12 (7.5%) | 12 (7.0%) | 4 (2.1%) | 16 (4.4%) | 28 (5.4%) |
| Not reported | 29 (18.1%) | 23 (13.4%) | 20 (10.6%) | 43 (11.9%) | 72 (13.8%) |
| Ethnicity |  |  |  |  |  |
| Hispanic or Latino | 32 (20.0%) | 46 (26.7%) | 35 (18.6%) | 81 (22.5%) | 113 (21.7%) |
| Not Hispanic or Latino | 119 (74.4%) | 117 (68.0%) | 142 (75.5%) | 259 (71.9%) | 378 (72.7%) |
| Not reported | 9 (5.6%) | 9 (5.2%) | 11 (5.9%) | 20 (5.6%) | 29 (5.6%) |
| Mean weight, kg | 88.17 ±25.05 | 90.30 ±25.62 | 90.86 ±23.63 | 90.60 ±24.56 | 89.85 ±24.71 |
| Body-mass index^b^ |  |  |  |  |  |
| Mean | 31.41±8.07 | 31.05±8.17 | 31.62±8.13 | 31.35±8.14 | 31.37±8.11 |
| ≥30 | 78 (48.8%) | 78 (45.3%) | 94 (50.0%) | 172 (47.8%) | 250 (48.1%) |
| Median days COVID-19 illness prior to baseline (Q1:Q3) | 5.0  (3.0:6.0) | 5.0  (3.0:7.0) | 5.0  (3.0:7.0) | 5.0  (3.0:7.0) | 5.0  (3.0:7.0) |
| Baseline viral load |  |  |  |  |  |
| Median (min:max), log_10_ copies/mL | 7.3  (6.3:8.4) | 7.4  (6.2:8.5) | 7.3  (6.1:8.0) | 7.3  (6.2:8.2) | 7.3  (6.2:8.2) |
| >10^4^ copies/mL | 155 (96.9%) | 168 (97.7%) | 183 (97.3%) | 351 (97.5%) | 506 (97.3%) |
| >10^6^ copies/mL | 127 (79.4%) | 132 (76.6%) | 147 (78.2%) | 279 (77.5%) | 406 (78.1%) |
| Mean C-reactive protein, mg/L | 62.952 ±64.97 | 64.486 ±124.01 | 49.485 ±46.12 | 56.738 ±92.54 | 58.623 ±85.10 |
| Mean neutrophils-lymphocyte ratio | 3.613±2.57 | 2.340±2.13 | 8.505±6.91 | 4.806±5.06 | 4.359±4.11 |
| Concomitant medications | 158 (98.8%) | 170 (98.8%) | 187 (99.5%) | 357 (99.2%) | 515 (99.0%) |
| Remdesivir | 96 (60.0%) | 103 (59.9%) | 113 (60.1%) | 216 (60.0%) | 312 (60.0%) |
| Systemic corticosteroids | 118 (73.8%) | 119 (69.2%) | 135 (71.8%) | 254 (70.6%) | 372 (71.5%) |
| Use of supplemental oxygen (Y) | 72 (45.0%) | 78 (45.3%) | 85 (45.2%) | 163 (45.3%) | 235 (45.2%) |
| Non-invasive ventilation or high flow oxygen devices | 1 (1.4%) | 0 | 0 | 0 | 1 (0.4%) |
| Supplemental oxygen^c^ | 71 (98.6%) | 78 (100%) | 85 (100%) | 163 (100%) | 234 (99.6%) |
| Immunocompromised (Y) | 40 (25.0%) | 42 (24.4%) | 50 (26.6%) | 92 (25.6%) | 132 (25.4%) |
| **Seropositive^a^** | **n=201** | **n=191** | **n=178** | **n=369** | **n=570** |
| Age |  |  |  |  |  |
| Median (range) | 64.0  (24–100) | 60  (20–94) | 61.0  (20–97) | 60.0  (20–97) | 62.0  (20–100) |
| ≥65 | 96 (47.8%) | 71 (37.2%) | 73 (41.0%) | 144 (39.0%) | 240 (42.1%) |
| Male sex | 112 (55.7%) | 104 (54.5%) | 100 (56.2%) | 204 (55.3%) | 316 (55.4%) |
| Race |  |  |  |  |  |
| White | 129 (64.2%) | 116 (60.7%) | 114 (64.0%) | 230 (62.3%) | 359 (63.0%) |
| Black or African American | 23 (11.4%) | 27 (14.1%) | 14 (7.9%) | 41 (11.1%) | 64 (11.2%) |
| Asian | 7 (3.5%) | 11 (5.8%) | 9 (5.1%) | 20 (5.4%) | 27 (4.7%) |
| American Indian or Alaska Native | 8 (4.0%) | 7 (3.7%) | 11 (6.2%) | 18 (4.9%) | 26 (4.6%) |
| Native Hawaiian or Pacific Islander | 0 | 0 | 1 (0.6%) | 1 (0.3%) | 1 (0.2%) |
| Unknown | 14 (7.0%) | 11 (5.8%) | 13 (7.3%) | 24 (6.5%) | 38 (6.7%) |
| Not reported | 20 (10.0%) | 19 (9.9%) | 16 (9.0%) | 35 (9.5%) | 55 (9.6%) |
| Ethnicity |  |  |  |  |  |
| Hispanic or Latino | 75 (37.3%) | 79 (41.4%) | 64 (36.0%) | 143 (38.8%) | 218 (38.2%) |
| Not Hispanic or Latino | 117 (58.2%) | 104 (54.5%) | 108 (60.7%) | 212 (57.5%) | 329 (57.7%) |
| Not reported | 9 (4.5%) | 8 (4.2%) | 6 (3.4%) | 14 (3.8%) | 23 (4.0%) |
| Mean weight, kg | 85.09 ±22.38 | 86.85 ±23.46 | 87.77 ±25.46 | 87.29 ±24.42 | 86.51 ±23.72 |
| Body mass index^b^ |  |  |  |  |  |
| Mean | 30.26±7.19 | 30.90±7.62 | 30.76±8.35 | 30.84±7.97 | 30.63±7.70 |
| ≥30 | 89 (44.3%) | 90 (47.1%) | 83 (46.6%) | 173 (46.9%) | 262 (46.0%) |
| Median days COVID-19 illness prior to baseline (Q1:Q3) | 6.0  (4.0:8.0) | 7.0  (5.0:8.0) | 7.0  (5.0:8.0) | 7.0  (5.0:8.0) | 6.0  (4.0:8.0) |
| Baseline viral load |  |  |  |  |  |
| Median (Q1:Q3), log_10_ copies/mL | 5.5  (4.4:6.6) | 5.4  (4.3:6.7) | 5.6  (4.4:6.9) | 5.6  (4.3:6.8) | 5.6  (4.4:6.7) |
| >10^4^ copies/mL | 172 (85.6%) | 156 (81.7%) | 145 (81.5%) | 301 (81.6%) | 473 (83.0%) |
| >10^6^ copies/mL | 80 (39.8%) | 73 (38.2%) | 72 (40.4%) | 145 (39.3%) | 225 (39.5%) |
| Mean C-reactive protein, mg/L | 87.252 ±71.19 | 85.378 ±69.29 | 94.742 ±110.15 | 89.726 ±90.56 | 88.844 ±84.11 |
| Mean neutrophils-lymphocyte ratio | 7.603±7.17 | 0 | 7.510±2.43 | 7.510±2.43 | 7.572±5.66 |
| Concomitant medications | 198 (98.5%) | 186 (97.4%) | 175 (98.3%) | 361 (97.8%) | 559 (98.1%) |
| Remdesivir | 102 (50.7%) | 87 (45.5%) | 93 (52.2%) | 180 (48.8%) | 282 (49.5%) |
| Systemic corticosteroids | 151 (75.1%) | 142 (74.3%) | 151 (84.8%) | 293 (79.4%) | 444 (77.9%) |
| Use of supplemental oxygen (Y) | 137 (68.2%) | 124 (64.9%) | 122 (68.5%) | 246 (66.7%) | 383 (67.2%) |
| Supplemental oxygen^c^ | 137 (100%) | 124 (100%) | 122 (100%) | 246 (100%) | 383 (100%) |
| Immunocompromised (Y) | 35 (17.4%) | 31 (16.2%) | 24 (13.5%) | 55 (14.9%) | 90 (15.8%) |
| **Other^a^** | **n=32** | **n=43** | **n=32** | **n=75** | **n=107** |
| Age |  |  |  |  |  |
| Median (range) | 65.5 (32–88) | 61.0 (26–89) | 60.0 (29–93) | 61.0 (26–93) | 62.0 (26–93) |
| ≥65 | 19 (59.4%) | 18 (41.9%) | 10 (31.3%) | 28 (37.3%) | 47 (43.9%) |
| Male sex | 16 (50.0%) | 21 (48.8%) | 15 (46.9%) | 36 (48.0%) | 52 (48.6%) |
| Race |  |  |  |  |  |
| White | 17 (53.1%) | 23 (53.5%) | 18 (56.3%) | 41 (54.7%) | 58 (54.2%) |
| Black or African American | 5 (15.6%) | 9 (20.9%) | 1 (3.1%) | 10 (13.3%) | 15 (14.0%) |
| Asian | 2 (6.3%) | 0 | 3 (9.4%) | 3 (4.0%) | 5 (4.7%) |
| American Indian or Alaska Native | 0 | 0 | 0 | 0 | 0 |
| Native Hawaiian or Pacific Islander | 0 | 0 | 0 | 0 | 0 |
| Unknown | 0 | 5 (11.6%) | 5 (15.6%) | 10 (13.3%) | 10 (9.3%) |
| Not reported | 8 (25.0%) | 6 (14.0%) | 5 (15.6%) | 11 (14.7%) | 19 (17.8%) |
| Ethnicity |  |  |  |  |  |
| Hispanic or Latino | 8 (25.0%) | 12 (27.9%) | 9 (28.1%) | 21 (28.0%) | 29 (27.1%) |
| Not Hispanic or Latino | 24 (75.0%) | 30 (69.8%) | 19 (59.4%) | 49 (65.3%) | 73 (68.2%) |
| Not reported | 0 | 1 (2.3%) | 4 (12.5%) | 5 (6.7%) | 5 (4.7%) |
| Mean weight, kg | 92.73±20.21 | 93.14±27.88 | 84.64±24.73 | 89.51±26.74 | 90.45±24.96 |
| Body mass index^b^ |  |  |  |  |  |
| Mean | 31.50±5.64 | 32.91±8.37 | 30.66±8.15 | 31.96±8.30 | 31.82±7.58 |
| ≥30 | 19 (59.4%) | 24 (55.8%) | 13 (40.6%) | 37 (49.3%) | 56 (52.3%) |
| Median days COVID-19 illness prior to baseline (Q1:Q3) | 6.0  (4.0:8.0) | 5.0  (3.0:7.0) | 5.0  (3.0:8.0) | 5.0  (3.0:7.0) | 5.0  (3.0:7.0) |
| Baseline viral load |  |  |  |  |  |
| Median (Q1:Q3), log_10_ copies/ml | 6.7 (3:10) | 6.5 (3:9) | 6.1 (3:9) | 6.3 (3:9) | 6.5 (3:10) |
| >10^4^ copies/mL | 29 (90.6%) | 42 (97.7%) | 31 (96.9%) | 73 (97.3%) | 102 (95.3%) |
| >10^6^ copies/mL | 22 (68.8%) | 26 (60.5%) | 17 (53.1%) | 43 (57.3%) | 65 (60.7%) |
| Mean C-reactive protein, mg/L | 58.053 ±55.41 | 60.443 ±71.54 | 72.257 ±68.11 | 65.269 ±69.91 | 63.176 ±65.84 |
| Mean neutrophils-lymphocyte ratio | 0 | 0 | 0 | 0 | 0 |
| Concomitant medications | 32 (100%) | 43 (100%) | 32 (100%) | 75 (100%) | 107 (100%) |
| Remdesivir | 22 (68.8%) | 22 (51.2%) | 19 (59.4%) | 41 (54.7%) | 63 (58.9%) |
| Systemic corticosteroids | 25 (78.1%) | 33 (76.7%) | 21 (65.6%) | 54 (72.0%) | 79 (73.8%) |
| Use of supplemental oxygen (Y) | 17 (53.1%) | 21 (48.8%) | 16 (50.0%) | 37 (49.3%) | 54 (50.5%) |
| Supplemental oxygen^c^ | 17 (100%) | 21 (100%) | 16 (100%) | 37 (100%) | 54 (100%) |
| Immunocompromised (Y) | 10 (31.3%) | 14 (32.6%) | 11 (34.4%) | 25 (33.3%) | 35 (32.7%) |

Data are n (%), median (IQR), or mean ±SD.

Abbreviations: CAS+IMD, casirivimab and imdevimab; COVID-19, coronavirus disease 2019; IQR, interquartile range; IV, intravenous; mFAS, modified full analysis set; SD, standard deviation.

^a^mFAS presented.

^b^The body-mass index is the weight in kilograms divided by the square of the height in meters.

^c^Not requiring high-flow oxygen devices.

Supplementary Table 3. Time to discharge for patients on low-flow or no supplemental oxygen

|  | **Placebo (n = 393)** | **CAS+IMD 2.4 g IV (n = 406)** | **CAS+IMD 8.0 g IV (n = 398)** | **CAS+IMD combined (n = 804)** |
| --- | --- | --- | --- | --- |
| Kaplan–Meier estimate |  |  |  |  |
| Number of patients assessed | 393 | 406 | 398 | 804 |
| Number of patients with events^a^ | 333 | 363 | 349 | 712 |
| Number of patients censored^a^ | 60 | 43 | 49 | 92 |
| 25% quantile (95% CI) | 3.0  (2.0, 3.0) | 2.0  (., .) | 2.0  (., .) | 2.0  (., .) |
| Median (95% CI)^b^ | 4.0  (4.0, 5.0) | 4.0  (4.0, 5.0) | 4.0  (4.0, 5.0) | 4.0  (4.0, 5.0) |
| 75% quantile (95% CI) | 11.0  (10.0, 15.0) | 9.0  (8.0, 10.0) | 10.0  (8.0, 12.0) | 9.0  (8.0, 10.0) |
| Min : Max^c^ | 0: 87+ | 0: 58+ | 0: 59+ | 0: 59+ |
| Hazard ratio versus placebo (95% CI) through day 29^d^ |  | 1.18  (1.01, 1.37) | 1.14  (0.98, 1.32) | 1.16  (1.01, 1.32) |
| Hazard ratio versus placebo (95% CI)^d^ |  | 1.16  (1.00, 1.35) | 1.13  (0.97, 1.32) | 1.15  (1.01, 1.31) |
| Rate ratio (95% CI)^e^ |  | 1.22  (1.04, 1.43) | 1.14  (0.97, 1.34) | 1.17  (1.03, 1.35) |

Abbreviations: CAS+IMD, casirivimab and imdevimab; CI, confidence interval; IV, intravenous.

^a^The event is the first discharge and alive. Time is days since first dose date until first discharge. Patients who did not experience the event and alive, time = (end of study date – date of first dose). If patients died, time = (168 days for phase I patients or 56 days for patients in other phases).

^b^Two-sided 95% CI is computed by Brookmeyer and Crowley method (log transformation)

^c^+ indicates censored observation.

^d^The hazard ratios and 95% CIs are estimated using Cox proportional hazard model with terms for treatment, baseline serostatus (positive, negative, and other), and the type of background standard-of-care (antiviral therapies and non-antiviral therapies) as fixed effect. Hazard ratio >1 implies CAS+IMD is better than placebo.

^e^The log-rank ‘observed minus expected’ statistic (and its variance) was used to calculate the 1-step estimate of the event rate ratio and CI.

Supplementary Table 4. Death or readmission at day 29 and day 57 for patients on low-flow or no supplemental oxygen

|  | **Placebo n/N (%)** | **CAS+IMD 2.4 g IV n/N (%)** | **CAS+IMD 8.0 g IV n/N (%)** | **CAS+IMD combined n/N (%)** |
| --- | --- | --- | --- | --- |
| **Seronegative mFAS** | **n = 160** | **n = 172** | **n = 188** | **n = 360** |
| Day 29 | 39/160 (24.4%) | 20/172 (11.6%) | 24/188 (12.8%) | 44/360 (12.2%) |
| 95% CI^a^ | 17.9–31.8 | 7.2–17.4 | 8.4–18.4 | 9.0–16.1 |
| Relative risk versus placebo (unstratified) |  | 0.477 | 0.524 | 0.501 |
| 95% CI^b^ |  | 0.291, 0.782 | 0.330, 0.832 | 0.340, 0.740 |
| Relative risk reduction versus placebo (unstratified) |  | 52.3% | 47.6% | 49.9% |
| 95% CI^b^ |  | 21.8–70.9% | 16.8–67.0% | 26.0–66.0% |
| Day 57 | 46/160 (28.8%) | 29/172 (16.9%) | 36/188 (19.1%) | 65/360 (18.1%) |
| 95% CI^a^ | 21.9–36.4 | 11.6–23.3 | 13.8–25.5 | 14.2–22.4 |
| Relative risk versus placebo (unstratified) |  | 0.586 | 0.666 | 0.628 |
| 95% CI^b^ |  | 0.388–0.885 | 0.455–0.976 | 0.452–0.872 |
| Relative risk reduction versus placebo (unstratified) |  | 41.4% | 33.4% | 37.2% |
| 95% CI^b^ |  | 11.5–61.2 | 2.4–54.5 | 12.8–54.8 |
| **Overall mFAS** | **N=393** | **N=406** | **N=393** | **N=804** |
| Day 29 | 67/393 (17.0%) | 44/406 (10.8%) | 59/398 (14.8%) | 103/804 (12.8%) |
| 95% CI^a^ | 13.5–21.1 | 8.0–14.3 | 11.5–18.7 | 10.6–15.3 |
| Relative risk versus placebo (unstratified) |  | 0.636 | 0.870 | 0.751 |
| 95% CI^b^ |  | 0.446–0.906 | 0.631–1.199 | 0.566–0.997 |
| Relative risk reduction versus placebo (unstratified) |  | 36.4% | 13.0% | 24.9% |
| 95% CI^b^ |  | 9.4–55.4 | ‑19.9–36.9 | 0.3–43.4 |
| Day 57 | 81/393 (20.6%) | 62/406 (15.3%) | 74/398 (18.6%) | 136/804 (16.9%) |
| 95% CI^a^ | 16.7–25.0 | 11.9–19.1 | 14.9–22.8 | 14.4–19.7 |
| Relative risk versus placebo (unstratified) |  | 0.741 | 0.902 | 0.821 |
| 95% CI^b^ |  | 0.549–1.000 | 0.680–1.197 | 0.641–1.051 |
| Relative risk reduction versus placebo (unstratified) |  | 25.9% | 9.8% | 17.9% |
| 95% CI^b^ |  | -0.0–45.1 | -19.7–32.0 | -5.1–35.9 |
| **High-viral load mFAS (>10^6^ copies/mL)** | **N=229** | **N=231** | **N=236** | **N=476** |
| Day 29 | 48/229 (21.0%) | 35/231 (15.2%) | 41/236 (17.4%) | 76/467 (16.3%) |
| 95% CI^a^ | 15.9–26.8 | 10.8–20.4 | 12.8–22.8 | 13.0–19.9 |
| Relative risk versus placebo (unstratified) |  | 0.723 | 0.829 | 0.776 |
| 95% CI^b^ |  | 0.487–1.073 | 0.570–1.206 | 0.561–1.074 |
| Relative risk reduction versus placebo (unstratified) |  | 27.7% | 17.1% | 22.4% |
| 95% CI |  | –7.3–51.3 | –20.6–43.0 | –7.4–43.9 |
| Day 57 | 59/229 (25.8%) | 47/231 (20.3%) | 54/236 (22.9%) | 101/467 (21.6%) |
| 95% CI^a^ | 20.2–31.9 | 15.3–26.1 | 17.7–28.8 | 18.0–25.6 |
| Relative risk versus placebo (unstratified) |  | 0.790 | 0.888 | 0.839 |
| 95% CI |  | 0.564–1.106 | 0.644–1.225 | 0.635–1.110 |
| Relative risk reduction versus placebo (unstratified) |  | 21.0% | 11.2% | 16.1% |
| 95% CI^b^ |  | -10.6–43.6 | -22.5–35.6 | -11.0–36.5 |

Data are n (%). N = number of patients in each treatment group; n = total number of patients who died or readmitted on or before day X; % = n/N.

Abbreviations: CAS+IMD, casirivimab and imdevimab; CI, confidence interval; IV, intravenous; mFAS, modified full analysis set.

^a^95% CIs were estimated using the exact Clopper-Pearson method.

^b^95% CIs for the relative risk and relative risk reduction (1 – relative risk) use the Farrington-Manning method.

**Supplementary Table 5. Adverse events leading to death in patients on low-flow oxygen**

|  | **Placebo (n = 469)** | **CAS+IMD  2.4 g IV (n = 470)** | **CAS+IMD  8.0 g IV (n = 471)** | **CAS+IMD  combined doses (n = 941)** |
| --- | --- | --- | --- | --- |
| Number of TEAEs leading to death | 72 | 49 | 61 | 110 |
| Number of patients with at least 1 TEAE leading to death | 72 (15.4%) | 49 (10.4%) | 59 (12.5%) | 108 (11.5%) |
| Respiratory, thoracic, and mediastinal disorders | 29 (6.2%) | 21 (4.5%) | 19 (4.0%) | 40 (4.3%) |
| Acute respiratory failure | 12 (2.6%) | 11 (2.3%) | 8 (1.7%) | 19 (2.0%) |
| Respiratory failure | 9 (1.9%) | 5 (1.1%) | 6 (1.3%) | 11 (1.2%) |
| Hypoxia | 4 (0.9%) | 0 | 3 (0.6%) | 3 (0.3%) |
| Pulmonary embolism | 1 (0.2%) | 3 (0.6%) | 0 | 3 (0.3%) |
| Acute respiratory distress syndrome | 1 (0.2%) | 1 (0.2%) | 1 (0.2%) | 2 (0.2%) |
| Chronic obstructive pulmonary disease | 0 | 0 | 1 (0.2%) | 1 (0.1%) |
| Pulmonary artery thrombosis | 0 | 1 (0.2%) | 0 | 1 (0.1%) |
| Idiopathic pulmonary fibrosis | 1 (0.2%) | 0 | 0 | 0 |
| Pneumonitis | 1 (0.2%) | 0 | 0 | 0 |
| Infections and infestations | 19 (4.1%) | 9 (1.9%) | 22 (4.7%) | 31 (3.3%) |
| COVID-19 | 6 (1.3%) | 4 (0.9%) | 10 (2.1%) | 14 (1.5%) |
| Septic shock | 3 (0.6%) | 2 (0.4%) | 5 (1.1%) | 7 (0.7%) |
| COVID-19 pneumonia | 5 (1.1%) | 2 (0.4%) | 4 (0.8%) | 6 (0.6%) |
| Sepsis | 0 | 1 (0.2%) | 1 (0.2%) | 2 (0.2%) |
| Device related bacteremia | 0 | 0 | 1 (0.2%) | 1 (0.1%) |
| Pneumonia | 1 (0.2%) | 0 | 1 (0.2%) | 1 (0.1%) |
| Pulmonary sepsis | 3 (0.6%) | 0 | 0 | 0 |
| Superinfection bacterial | 1 (0.2%) | 0 | 0 | 0 |
| Cardiac disorders | 8 (1.7%) | 9 (1.9%) | 7 (1.5%) | 16 (1.7%) |
| Cardiac arrest | 1 (0.2%) | 4 (0.9%) | 1 (0.2%) | 5 (0.5%) |
| Cardio-respiratory arrest | 3 (0.6%) | 2 (0.4%) | 2 (0.4%) | 4 (0.4%) |
| Acute myocardial infarction | 0 | 0 | 2 (0.4%) | 2 (0.2%) |
| Cardiogenic shock | 0 | 1 (0.2%) | 1 (0.2%) | 2 (0.2%) |
| Bradycardia | 0 | 1 (0.2%) | 0 | 1 (0.1%) |
| Cardiac failure congestive | 0 | 0 | 1 (0.2%) | 1 (0.1%) |
| Ventricular tachycardia | 0 | 1 (0.2%) | 0 | 1 (0.1%) |
| Acute left ventricular failure | 1 (0.2%) | 0 | 0 | 0 |
| Atrial fibrillation | 1 (0.2%) | 0 | 0 | 0 |
| Cardiac failure | 1 (0.2%) | 0 | 0 | 0 |
| Pulseless electrical activity | 1 (0.2%) | 0 | 0 | 0 |
| General disorders and administration-site conditions | 7 (1.5%) | 5 (1.1%) | 9 (1.9%) | 14 (1.5%) |
| Multiple organ dysfunction syndrome | 4 (0.9%) | 2 (0.4%) | 7 (1.5%) | 9 (1.0%) |
| Death | 3 (0.6%) | 3 (0.6%) | 1 (0.2%) | 4 (0.4%) |
| Sudden cardiac death | 0 | 0 | 1 (0.2%) | 1 (0.1%) |
| Nervous system disorders | 2 (0.4%) | 2 (0.4%) | 0 | 2 (0.2%) |
| Encephalopathy | 0 | 1 (0.2%) | 0 | 1 (0.1%) |
| Metabolic encephalopathy | 0 | 1 (0.2%) | 0 | 1 (0.1%) |
| Cerebrovascular accident | 1 (0.2%) | 0 | 0 | 0 |
| Dementia Alzheimer’s type | 1 (0.2%) | 0 | 0 | 0 |
| Renal and urinary disorders | 1 (0.2%) | 1 (0.2%) | 1 (0.2%) | 2 (0.2%) |
| Acute kidney injury | 0 | 1 (0.2%) | 1 (0.2%) | 2 (0.2%) |
| Renal failure | 1 (0.2%) | 0 | 0 | 0 |
| Vascular disorders | 2 (0.4%) | 1 (0.2%) | 1 (0.2%) | 2 (0.2%) |
| Arteriosclerosis | 0 | 0 | 1 (0.2%) | 1 (0.1%) |
| Distributive shock | 0 | 1 (0.2%) | 0 | 1 (0.1%) |
| Shock | 2 (0.4%) | 0 | 0 | 0 |
| Gastrointestinal disorders | 2 (0.4%) | 0 | 1 (0.2%) | 1 (0.1%) |
| Retroperitoneal hemorrhage | 0 | 0 | 1 (0.2%) | 1 (0.1%) |
| Gastrointestinal hemorrhage | 1 (0.2%) | 0 | 0 | 0 |
| Retroperitoneal hematoma | 1 (0.2%) | 0 | 0 | 0 |
| Hepatobiliary disorders | 1 (0.2%) | 1 (0.2%) | 0 | 1 (0.1%) |
| Hepatic cirrhosis | 0 | 1 (0.2%) | 0 | 1 (0.1%) |
| Acute hepatic failure | 1 (0.2%) | 0 | 0 | 0 |
| Investigations | 0 | 0 | 1 (0.2%) | 1 (0.1%) |
| Oxygen saturation decreased | 0 | 0 | 1 (0.2%) | 1 (0.1%) |
| Injury, poisoning, and procedural complications | 1 (0.2%) | 0 | 0 | 0 |
| Brain herniation | 1 (0.2%) | 0 | 0 | 0 |

Data are n (%). Primary SOCs are sorted according to decreasing order of frequency of the combined treatment group. Within each SOC, Preferred Terms are sorted by decreasing frequency. A patient who reported 2 or more adverse events with different preferred terms within the same system organ class is counted only once in that system organ class. A patient who reported 2 or more adverse events with the same preferred term is counted only once for that term.

Abbreviations: CAS+IMD, casirivimab and imdevimab; IV, intravenous; SOC, System Organ Class; TEAE, treatment-emergent adverse event.

**Supplementary Table 6. Adverse events leading to death in patients on no supplemental oxygen**

|  | **Placebo (n = 198)** | **CAS+IMD  2.4 g IV (n = 202)** | **CAS+IMD  8.0 g IV (n = 197)** | **CAS+IMD  combined doses (n = 399)** |
| --- | --- | --- | --- | --- |
| Number of TEAEs leading to death | 15 | 8 | 7 | 15 |
| Number of patients with at least 1 TEAE leading to death | 15 (7.6%) | 8 (4.0%) | 7 (3.6%) | 15 (3.8%) |
| Infections and infestations | 6 (3.0%) | 1 (0.5%) | 4 (2.0%) | 5 (1.3%) |
| COVID-19 | 5 (2.5%) | 0 | 3 (1.5%) | 3 (0.8%) |
| Urosepsis | 0 | 1 (0.5%) | 0 | 1 (0.3%) |
| Viral myocarditis | 0 | 0 | 1 (0.5%) | 1 (0.3%) |
| COVID-19 pneumonia | 1 (0.5%) | 0 | 0 | 0 |
| Respiratory, thoracic, and mediastinal disorders | 3 (1.5%) | 2 (1.0%) | 1 (0.5%) | 3 (0.8%) |
| Acute respiratory failure | 1 (0.5%) | 1 (0.5%) | 1 (0.5%) | 2 (0.5%) |
| Respiratory failure | 1 (0.5%) | 1 (0.5%) | 0 | 1 (0.3%) |
| Alveolar lung disease | 1 (0.5%) | 0 | 0 | 0 |
| General disorders and administration-site conditions | 1 (0.5%) | 2 (1.0%) | 0 | 2 (0.5%) |
| Death | 1 (0.5%) | 2 (1.0%) | 0 | 2 (0.5%) |
| Metabolism and nutrition disorders | 0 | 0 | 1 (0.5%) | 1 (0.3%) |
| Failure to thrive | 0 | 0 | 1 (0.5%) | 1 (0.3%) |
| Neoplasms benign,­ malignant, and unspecified (including cysts and polyps) | 0 | 1 (0.5%) | 0 | 1 (0.3%) |
| Colon cancer | 0 | 1 (0.5%) | 0 | 1 (0.3%) |
| Nervous system disorders | 1 (0.5%) | 0 | 1 (0.5%) | 1 (0.3%) |
| Hemorrhage intracranial | 0 | 0 | 1 (0.5%) | 1 (0.3%) |
| Cerebrovascular accident | 1 (0.5%) | 0 | 0 | 0 |
| Psychiatric disorders | 0 | 1 (0.5%) | 0 | 1 (0.3%) |
| Delirium | 0 | 1 (0.5%) | 0 | 1 (0.3%) |
| Vascular disorders | 0 | 1 (0.5%) | 0 | 1 (0.3%) |
| Shock hemorrhagic | 0 | 1 (0.5%) | 0 | 1 (0.3%) |
| Blood and lymphatic system disorders | 1 (0.5%) | 0 | 0 | 0 |
| Sickle cell anemia with crisis | 1 (0.5%) | 0 | 0 | 0 |
| Cardiac disorders | 2 (1.0%) | 0 | 0 | 0 |
| Cardiac arrest | 2 (1.0%) | 0 | 0 | 0 |
| Renal and urinary disorders | 1 (0.5%) | 0 | 0 | 0 |
| End stage renal disease | 1 (0.5%) | 0 | 0 | 0 |

Data are n (%). Primary SOCs are sorted according to decreasing order of frequency of the combined treatment group. Within each SOC, Preferred Terms are sorted by decreasing frequency. A patient who reported 2 or more adverse events with different preferred terms within the same system organ class is counted only once in that system organ class. A patient who reported 2 or more adverse events with the same preferred term is counted only once for that term.

Abbreviations: CAS+IMD, casirivimab and imdevimab; IV, intravenous; SOC, System Organ Class; TEAE, treatment-emergent adverse event.

**Supplementary Table 7. Adverse events of special interest in patients on low-flow oxygen**

|  | **Placebo (n = 469)** | **CAS+IMD  2.4 g IV (n = 470)** | **CAS+IMD  8.0 g IV (n = 471)** | **CAS+IMD  combined doses (n = 941)** |
| --- | --- | --- | --- | --- |
| Number of AESIs | 9 | 12 | 21 | 33 |
| Number of patients with at least 1 AESI | 6 (1.3%) | 10 (2.1%) | 14 (3.0%) | 24 (2.6%) |
| Grade ≥2 infusion-related reactions through Day 4 | 5 (1.1%) | 7 (1.5%) | 11 (2.3%) | 18 (1.9%) |
| Hypoxia | 1 (0.2%) | 0 | 3 (0.6%) | 3 (0.3%) |
| Chest pain | 0 | 1 (0.2%) | 1 (0.2%) | 2 (0.2%) |
| Pyrexia | 0 | 1 (0.2%) | 1 (0.2%) | 2 (0.2%) |
| Acute respiratory failure | 0 | 1 (0.2%) | 0 | 1 (0.1%) |
| Anaphylactic reaction | 0 | 0 | 1 (0.2%) | 1 (0.1%) |
| Chills | 1 (0.2%) | 0 | 1 (0.2%) | 1 (0.1%) |
| Dyspnea | 0 | 0 | 1 (0.2%) | 1 (0.1%) |
| Hemoptysis | 0 | 0 | 1 (0.2%) | 1 (0.1%) |
| Hyperkalemia | 0 | 0 | 1 (0.2%) | 1 (0.1%) |
| Hypoesthesia | 0 | 1 (0.2%) | 0 | 1 (0.1%) |
| Infusion-related reaction | 1 (0.2%) | 0 | 1 (0.2%) | 1 (0.1%) |
| Infusion-site pain | 0 | 0 | 1 (0.2%) | 1 (0.1%) |
| Paresthesia | 0 | 1 (0.2%) | 0 | 1 (0.1%) |
| Pruritus | 0 | 1 (0.2%) | 0 | 1 (0.1%) |
| Systemic inflammatory response syndrome | 0 | 1 (0.2%) | 0 | 1 (0.1%) |
| Tachypnoea | 0 | 1 (0.2%) | 0 | 1 (0.1%) |
| Alanine aminotransferase increased | 1 (0.2%) | 0 | 0 | 0 |
| Aspartate aminotransferase increased | 1 (0.2%) | 0 | 0 | 0 |
| Urticaria | 1 (0.2%) | 0 | 0 | 0 |
| Grade ≥2 hypersensitivity reactions through day 29 | 1 (0.2%) | 3 (0.6%) | 4 (0.8%) | 7 (0.7%) |
| Pulmonary embolism | 0 | 2 (0.4%) | 0 | 2 (0.2%) |
| Anxiety | 0 | 0 | 1 (0.2%) | 1 (0.1%) |
| Aspartate aminotransferase increased | 0 | 1 (0.2%) | 0 | 1 (0.1%) |
| Chills | 0 | 0 | 1 (0.2%) | 1 (0.1%) |
| Deep vein thrombosis | 0 | 1 (0.2%) | 0 | 1 (0.1%) |
| Dyspnea | 0 | 0 | 1 (0.2%) | 1 (0.1%) |
| Flushing | 0 | 0 | 1 (0.2%) | 1 (0.1%) |
| Headache | 0 | 0 | 1 (0.2%) | 1 (0.1%) |
| Hypersensitivity | 0 | 0 | 1 (0.2%) | 1 (0.1%) |
| Nausea | 0 | 0 | 1 (0.2%) | 1 (0.1%) |
| Edema | 0 | 0 | 1 (0.2%) | 1 (0.1%) |
| Vomiting | 0 | 0 | 1 (0.2%) | 1 (0.1%) |
| Acute kidney injury | 1 (0.2%) | 0 | 0 | 0 |
| Hypotension | 1 (0.2%) | 0 | 0 | 0 |
| Hypoxia | 1 (0.2%) | 0 | 0 | 0 |

Data are n (%). Primary SOCs are sorted according to decreasing order of frequency of the combined treatment group. Within each SOC, Preferred Terms are sorted by decreasing frequency. A patient who reported 2 or more adverse events with different preferred terms within the same system organ class is counted only once in that system organ class. A patient who reported 2 or more adverse events with the same preferred term is counted only once for that term.

Abbreviations: AESI, adverse event of special interest; CAS+IMD, casirivimab and imdevimab; IV, intravenous; SOC, System Organ Class.

**Supplementary Table 8. Adverse events of special interest in patients on no supplemental oxygen**

|  | **Placebo (n = 198)** | **CAS+IMD  2.4 g IV (n = 202)** | **CAS+IMD  8.0 g IV (n = 197)** | **CAS+IMD  combined doses (n = 399)** |
| --- | --- | --- | --- | --- |
| Number of AESIs | 2 | 5 | 8 | 13 |
| Number of patients with at least 1 AESI | 2 (1.0%) | 4 (2.0%) | 6 (3.0%) | 10 (2.5%) |
| Grade ≥2 infusion-related reactions through Day 4 | 1 (0.5%) | 4 (2.0%) | 4 (2.0%) | 8 (2.0%) |
| Hypoxia | 0 | 3 (1.5%) | 0 | 3 (0.8%) |
| Tachycardia | 0 | 0 | 2 (1.0%) | 2 (0.5%) |
| Chills | 0 | 0 | 1 (0.5%) | 1 (0.3%) |
| Infusion-related reaction | 1 (0.5%) | 1 (0.5%) | 0 | 1 (0.3%) |
| Infusion-site extravasation | 0 | 1 (0.5%) | 0 | 1 (0.3%) |
| Pruritus | 0 | 0 | 1 (0.5%) | 1 (0.3%) |
| Grade ≥2 hypersensitivity reactions through Day 29 | 1 (0.5%) | 0 | 2 (1.0%) | 2 (0.5%) |
| Dizziness | 0 | 0 | 1 (0.5%) | 2 (0.5%) |
| Dyspnea | 0 | 0 | 1 (0.5%) | 1 (0.3%) |
| Hypoxia | 0 | 0 | 1 (0.5%) | 1 (0.3%) |
| Tachypnoea | 0 | 0 | 1 (0.5%) | 1 (0.3%) |
| Inflammatory marker increase | 1 (0.5%) | 0 | 0 | 0 |

Data are n (%). Primary SOCs are sorted according to decreasing order of frequency of the combined treatment group. Within each SOC, Preferred Terms are sorted by decreasing frequency. A patient who reported 2 or more adverse events with different preferred terms within the same system organ class is counted only once in that system organ class. A patient who reported 2 or more adverse events with the same preferred term is counted only once for that term.

Abbreviations: AESI, adverse event of special interest; CAS+IMD, casirivimab and imdevimab; IV, intravenous; SOC, System Organ Class.

Supplementary Table 9. Adverse events leading to death in patients on high-intensity oxygen

|  | **Placebo (n = 51)** | **CAS+IMD IV** | | |
| --- | --- | --- | --- | --- |
|  |  | **2.4 g  (n = 56)** | **8.0 g  (n = 54)** | **Combined (n = 110)** |
| Number of TEAEs leading to death | 13 | 25 | 19 | 44 |
| Number of patients with at least 1 TEAE leading to death | 13 (25.5%) | 25 (44.6%) | 19 (35.2%) | 44 (40.0%) |
| Infections and infestations | 6 (11.8%) | 9 (16.1%) | 10 (18.5%) | 19 (17.3%) |
| COVID-19 | 2 (3.9%) | 6 (10.7%) | 7 (13.0%) | 13 (11.8%) |
| COVID-19 pneumonia | 3 (5.9%) | 2 (3.6%) | 3 (5.6%) | 5 (4.5%) |
| Septic shock | 1 (2.0%) | 1 (1.8%) | 0 | 1 (0.9%) |
| Respiratory, thoracic, and mediastinal disorders | 5 (9.8%) | 13 (23.2%) | 4 (7.4%) | 17 (15.5%) |
| Acute respiratory failure | 0 | 7 (12.5%) | 3 (5.6%) | 10 (9.1%) |
| Respiratory failure | 4 (7.8%) | 3 (5.4%) | 1 (1.9%) | 4 (3.6%) |
| Acute respiratory distress syndrome | 0 | 1 (1.8%) | 0 | 1 (0.9%) |
| Hypoxia | 1 (2.0%) | 1 (1.8%) | 0 | 1 (0.9%) |
| Respiratory distress | 0 | 1 (1.8%) | 0 | 1 (0.9%) |
| Cardiac disorders | 1 (2.0%) | 2 (3.6%) | 3 (5.6%) | 5 (4.5%) |
| Cardiac arrest | 1 (2.0%) | 2 (3.6%) | 2 (3.7%) | 4 (3.6%) |
| Cardiac disorders |  |  |  |  |
| Bradycardia | 0 | 0 | 1 (1.9%) | 1 (0.9%) |
| General disorders and administration-site conditions | 0 | 1 (1.8%) | 1 (1.9%) | 2 (1.8%) |
| Multiple organ dysfunction syndrome | 0 | 1 (1.8%) | 1 (1.9%) | 2 (1.8%) |
| Nervous system disorders | 0 | 0 | 1 (1.9%) | 1 (0.9%) |
| Cerebrovascular accident | 0 | 0 | 1 (1.9%) | 1 (0.9%) |
| Gastrointestinal disorders | 1 (2.0%) | 0 | 0 | 0 |
| Intestinal ischemia | 1 (2.0%) | 0 | 0 | 0 |

Data are n (%). Results are presented from cohort 2 (phase II) prior to IDMC recommendation to pause enrolment into this cohort October 30, 2020. Primary SOCs are sorted according to decreasing order of frequency of the combined treatment group. Within each SOC, Preferred Terms are sorted by decreasing frequency. A patient who reported 2 or more adverse events with different preferred terms within the same system organ class is counted only once in that system organ class. A patient who reported 2 or more adverse events with the same preferred term is counted only once for that term.

Abbreviations: CAS+IMD, casirivimab and imdevimab; IDMC, independent data monitoring committee; IV, intravenous; SOC, System Organ Class.

Supplementary Table 10. Adverse events leading to death in patients on mechanical ventilation

|  | **Placebo (n = 12)** | **CAS+IMD IV** | | |
| --- | --- | --- | --- | --- |
|  |  | **2.4 g  (n = 12)** | **8.0 g  (n = 11)** | **Combined (n = 23)** |
| Number of TEAEs leading to death | 7 | 8 | 5 | 13 |
| Number of patients with at least 1 TEAE leading to death | 7 (58.3%) | 8 (66.7%) | 4 (36.4%) | 12 (52.2%) |
| Respiratory, thoracic, and mediastinal disorders | 4 (33.3%) | 4 (33.3%) | 1 (9.1%) | 5 (21.7%) |
| Respiratory failure | 1 (8.3%) | 3 (25.0%) | 0 | 3 (13.0%) |
| Acute respiratory failure | 2 (16.7%) | 1 (8.3%) | 1 (9.1%) | 2 (8.7%) |
| Respiratory arrest | 1 (8.3%) | 0 | 0 | 0 |
| Infections and infestations | 2 (16.7%) | 2 (16.7%) | 2 (18.2%) | 4 (17.4%) |
| COVID-19 | 2 (16.7%) | 1 (8.3%) | 2 (18.2%) | 3 (13.0%) |
| Viral cardiomyopathy | 0 | 1 (8.3%) | 0 | 1 (4.3%) |
| Cardiac disorders | 0 | 1 (8.3%) | 1 (9.1%) | 2 (8.7%) |
| Cardiac arrest | 0 | 1 (8.3%) | 1 (9.1%) | 2 (8.7%) |
| Renal and urinary disorders | 0 | 0 | 1 (9.1%) | 1 (4.3%) |
| Renal impairment | 0 | 0 | 1 (9.1%) | 1 (4.3%) |
| Vascular disorders | 0 | 1 (8.3%) | 0 | 1 (4.3%) |
| Hemorrhage | 0 | 1 (8.3%) | 0 | 1 (4.3%) |
| General disorders and administration-site conditions | 1 (8.3%) | 0 | 0 | 0 |
| Sudden cardiac death | 1 (8.3%) | 0 | 0 | 0 |

Data are n (%). Results are presented from cohort 3 (phase II) prior to IDMC recommendation to pause enrolment into this cohort October 30, 2020. Primary SOCs are sorted according to decreasing order of frequency of the combined treatment group. Within each SOC, Preferred Terms are sorted by decreasing frequency. A patient who reported 2 or more adverse events with different preferred terms within the same system organ class is counted only once in that system organ class. A patient who reported 2 or more adverse events with the same preferred term is counted only once for that term.

Abbreviations: CAS+IMD, casirivimab and imdevimab; IDMC, independent data monitoring committee; IV, intravenous; SOC, System Organ Class; TEAE, treatment-emergent adverse event.

Supplementary Figure 1. Schematic overview of the phase II/III study design

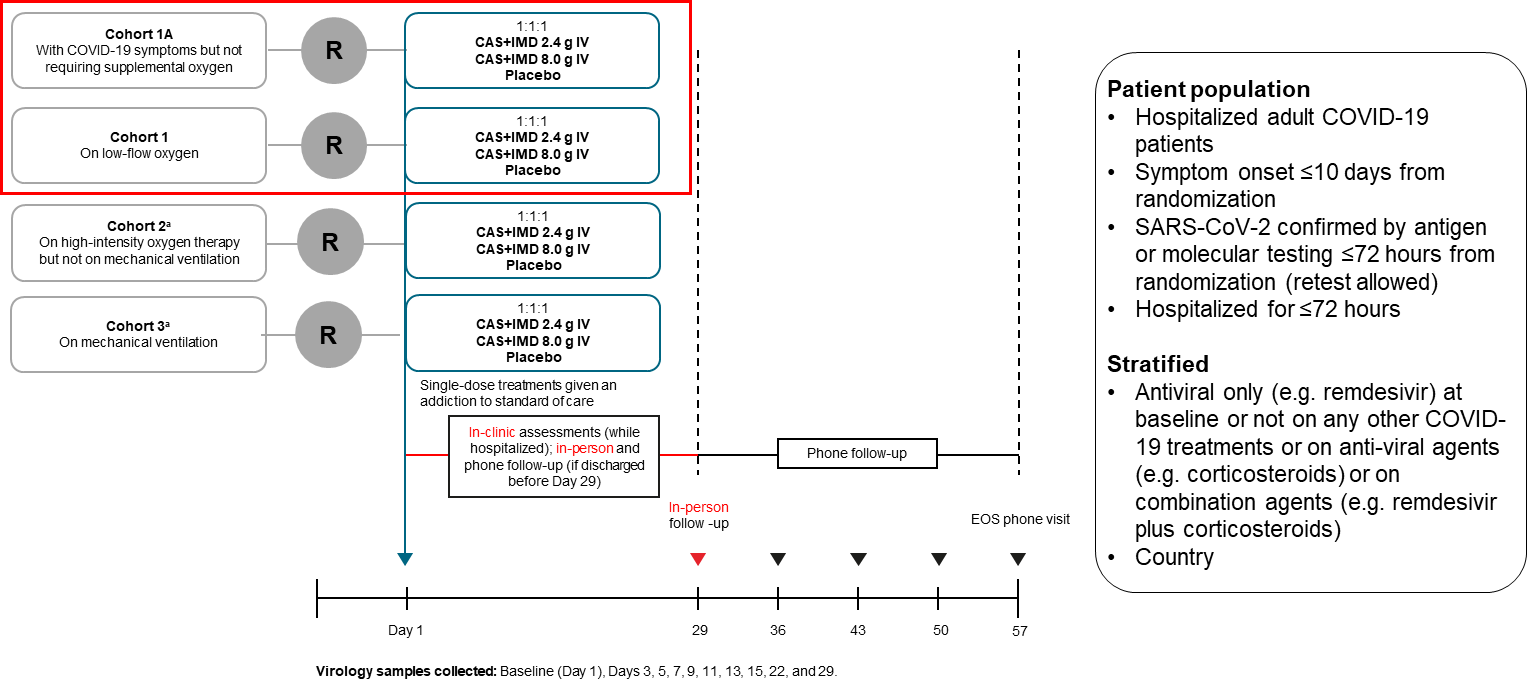

A schematic overview of is depicted for the phase II/III portion of the study design from day 1 through 57. The study was designed with 4 cohorts based on respiratory status at baseline. Only cohorts 1 and 1A (no supplemental oxygen and low-flow oxygen) are included in analyses; enrolment cohorts 2 and 3 (high-intensity oxygen and mechanical ventilation) was placed on hold per IDMC recommendation and not resumed prior to premature termination of the study for strategic reasons not related to safety.

Abbreviations: CAS+IMD, casirivimab and imdevimab; COVID-19, coronavirus disease 2019; EOS, end of study; IDMC, independent data monitoring committee; IV, intravenous; SARS-CoV-2, severe acute respiratory syndrome coronavirus 2;

^a^Per IDMC recommendation received on October 30, November 18, and December 10, 2020, patient enrolment in cohort 2 and cohort 3 was placed on hold (pending IDMC review of further data on patients who were enrolled in these cohorts) while it was recommended to proceed with enrolment of patients in cohort 1A and cohort 1. On April 9, 2021, Regeneron made a business decision to terminate patient enrolment in all cohorts of this study due to low recruitment.

Supplementary Figure 2. Flow diagram for the phase II/III population receiving low-flow or no supplemental oxygen

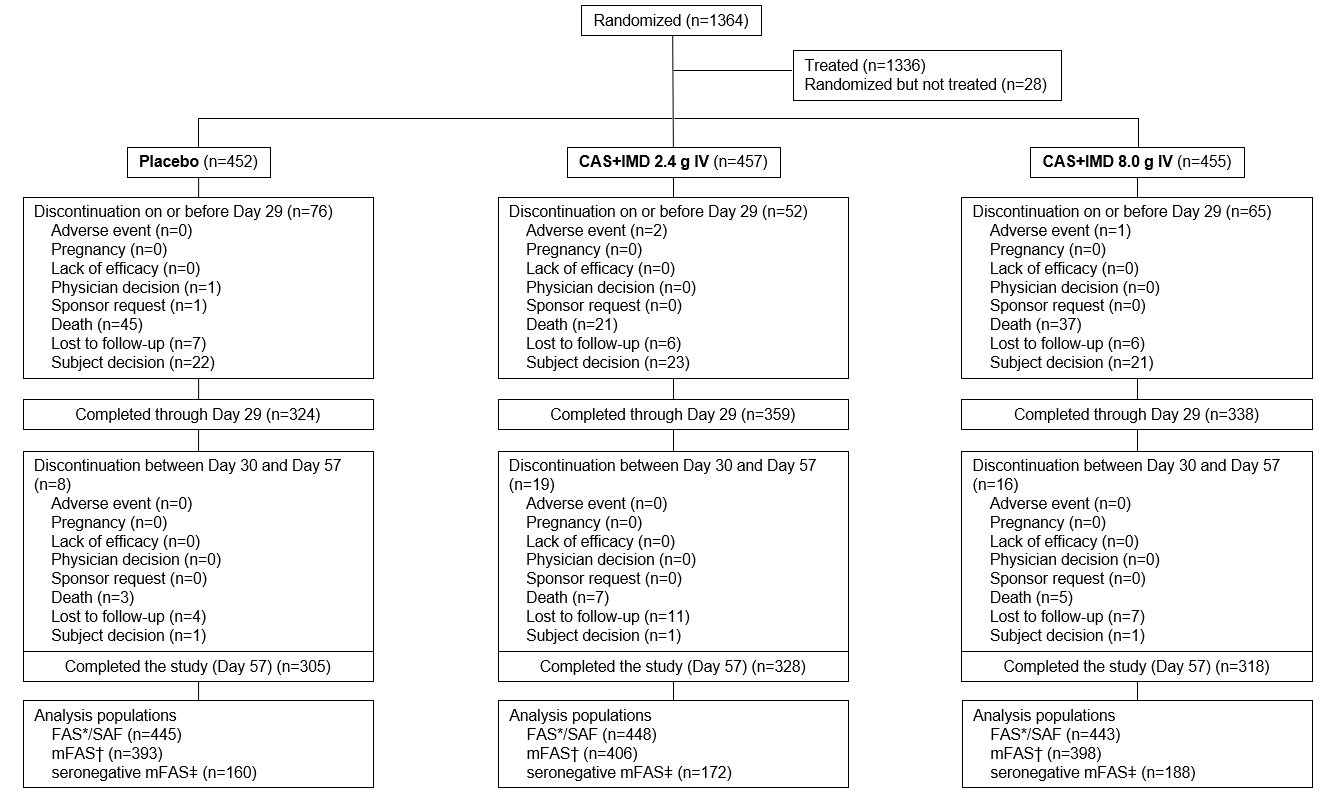

A flow diagram depicts patients randomized, treated, and discontinued for patients receiving either 2.4 or 8.0 g of CAS+IMD, or placebo.

Abbreviations: FAS, full analysis set; IV, intravenous; mFAS, modified full analysis set; RT-qPCR, quantitative reverse transcription polymerase chain reaction; SAF, safety analysis set; SARS-CoV-2, severe acute respiratory syndrome coronavirus 2.

*The FAS includes all randomized patients who received at least 1 dose (full or partial) of the study drug. Analysis of the FAS population will be done according to the treatment allocated (as randomized). The FAS is the same as the SAF for this study.

^†^The mFAS includes all FAS patients with a positive SARS-CoV-2 RT-qPCR conducted in the central laboratory in nasopharyngeal swab samples at randomization, and analysis is based on the treatment allocated (as randomized).

^‡^The seronegative mFAS is defined as all patients in mFAS with documented seronegative status at baseline.

Supplementary Figure 3. Viral load in the overall population from day 1 though day 29

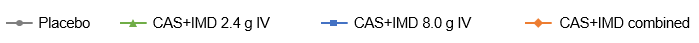

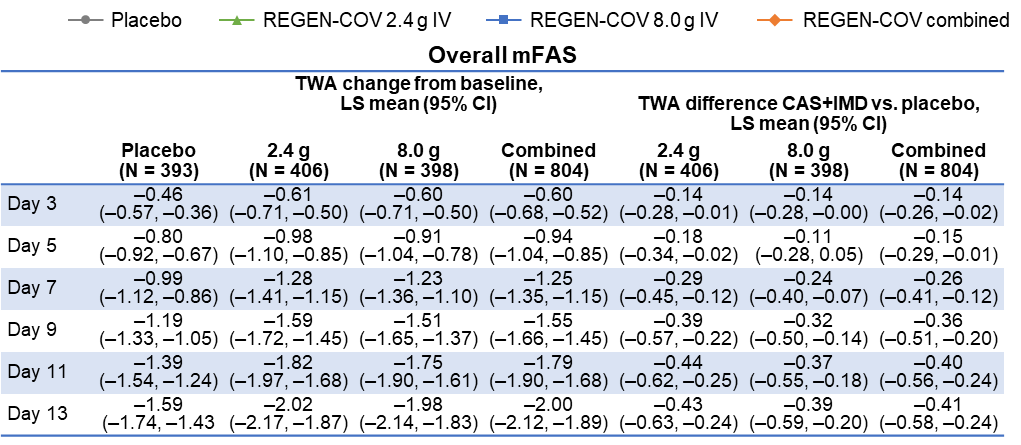

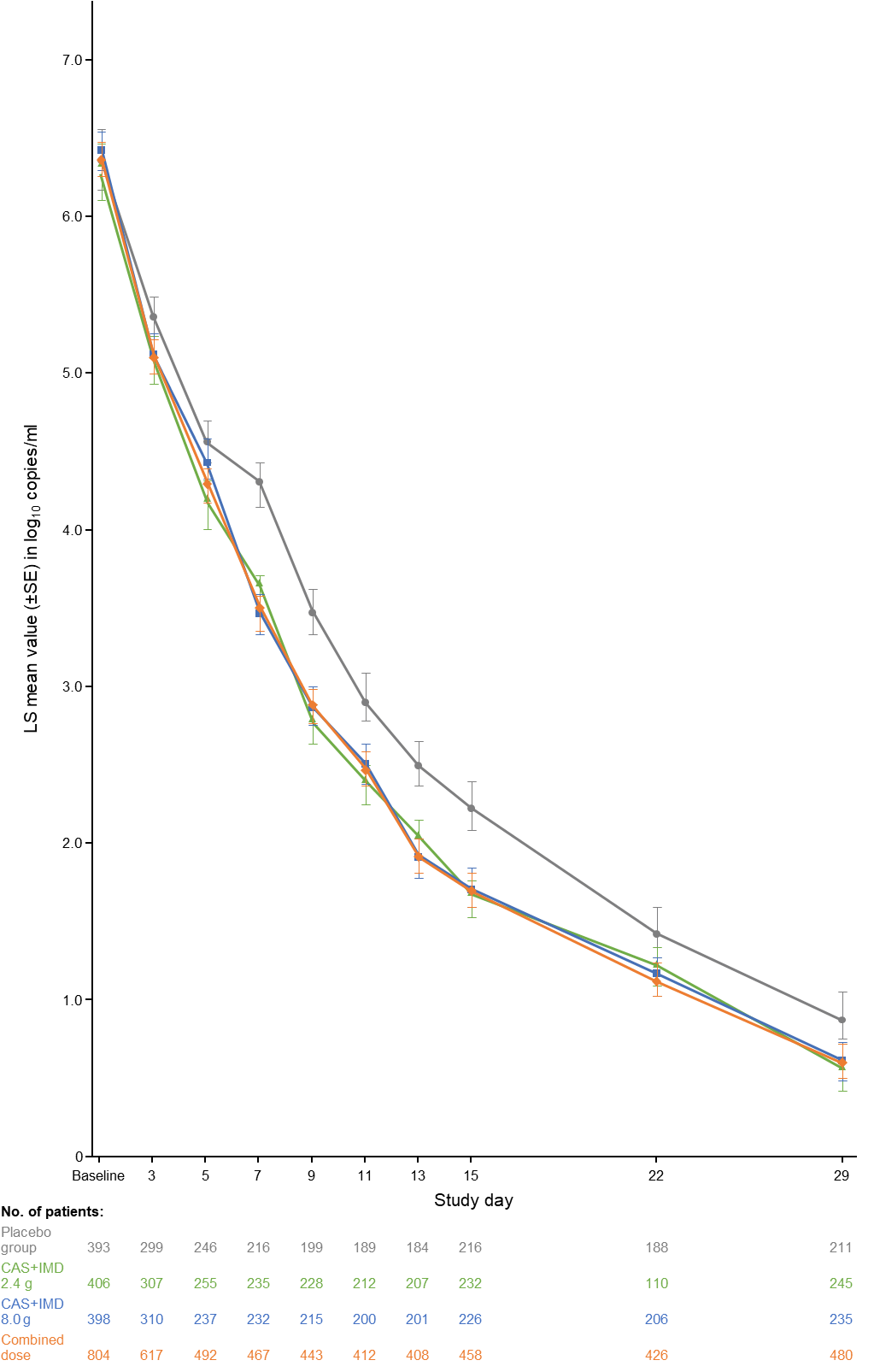

The graph displays LS mean viral load following administration of CAS+IMD (2.4 g, 8.0 g, or combined analysis of 2.4 and 8.0 g) or placebo in the overall population (seronegative, seropositive, and undetermined serostatus combined) by study day. The LLOQ is 2.85 log_10_ copies/mL.

Abbreviations: CAS+IMD, casirivimab and imdevimab; CI, 95% confidence-interval; IV, intravenous; LLOQ, lower limit of quantification; LS, least-squares; mFAS, modified full analysis set; PBO, placebo; SE, standard error; TWA, time-weighted average.

Supplementary Figure 4. Efficacy outcomes by serostatus and by dose from day 1 though day 29

A. **Low dose (2.4 g)**

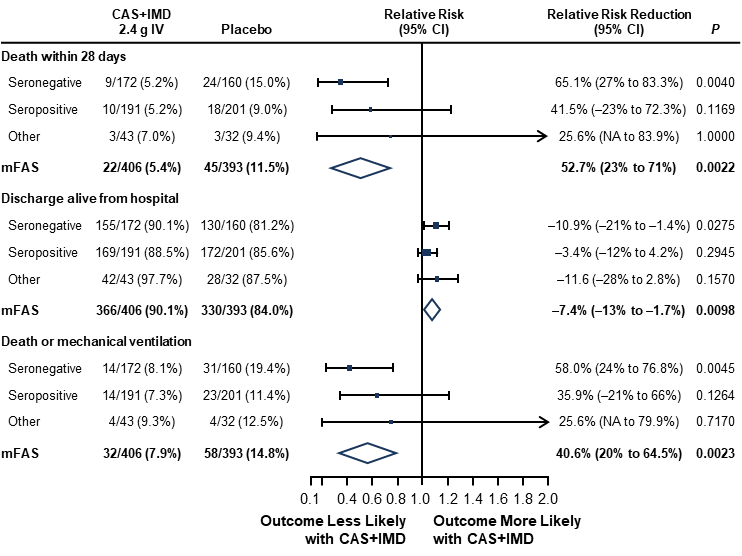

B. **High dose (8.0 g)**

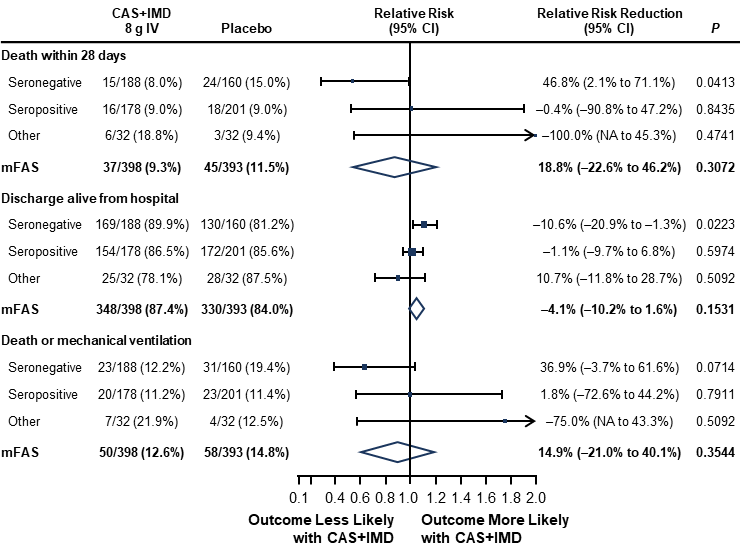

Panel A shows a forest plot of relative risk and relative risk reduction with 95% CI for CAS+IMD low-dose analysis (2.4 g) versus placebo. Parameters examined include death within 28 days, discharge alive from hospital, and death or mechanical ventilation. Populations analyzed include patients who tested negative for all SARS-CoV-2 antibodies at baseline (seronegative mFAS) patients who tested positive for any SARS-CoV-2 antibody at baseline (seropositive mFAS), those with inconclusive or missing baseline serology (other), and the overall population regardless of serostatus (overall mFAS). *P* values are considered nominal. Panel B shows a forest plot similar to Panel A, but for CAS+IMD high dose analysis (8.0 g) versus placebo.

Abbreviations: CAS+IMD, casirivimab and imdevimab; CI, confidence interval; IV, intravenous; mFAS, modified full analysis set; SARS-CoV-2, severe acute respiratory syndrome coronavirus 2.

Supplementary Figure 5. Separate efficacy outcomes for patient cohorts on low-flow or no supplemental oxygen by serostatus and by dose from day 1 though day 29

**A. Low-flow oxygen (cohort 1, phase III) combined analysis of 2.4 g and 8.0 g CAS+IMD**

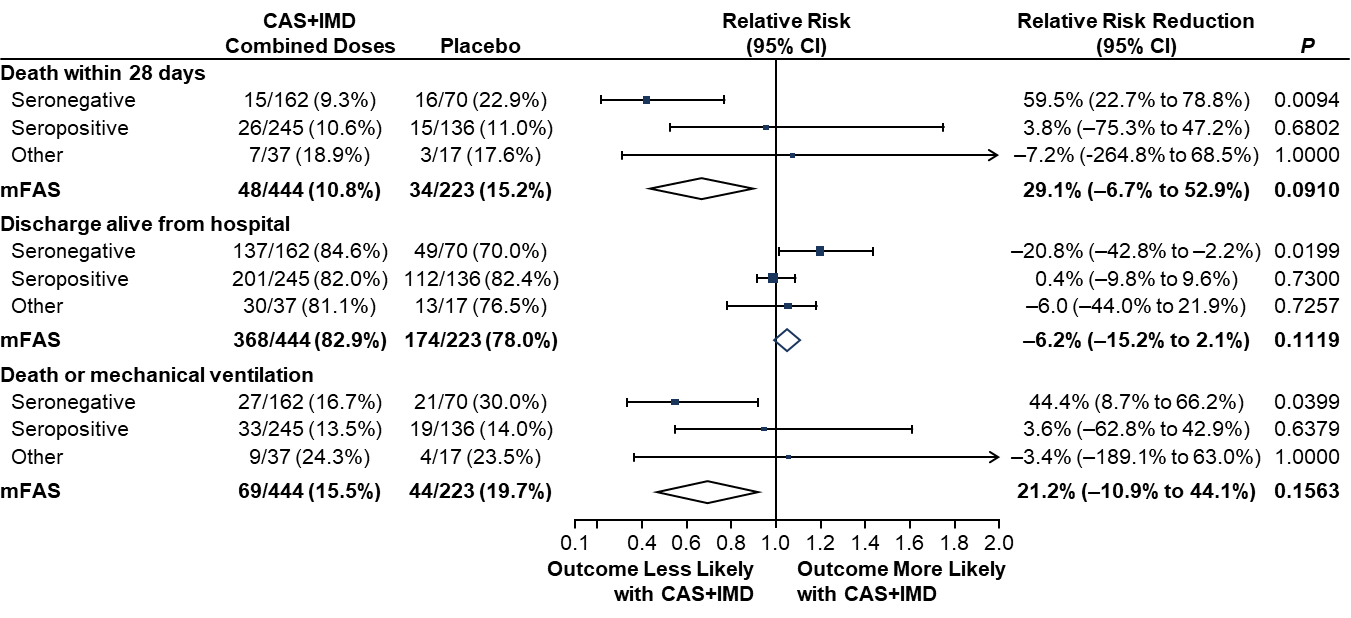

**B. No supplemental oxygen (cohort 1A, phase II) combined analysis of 2.4 g and 8.0 g CAS+IMD**

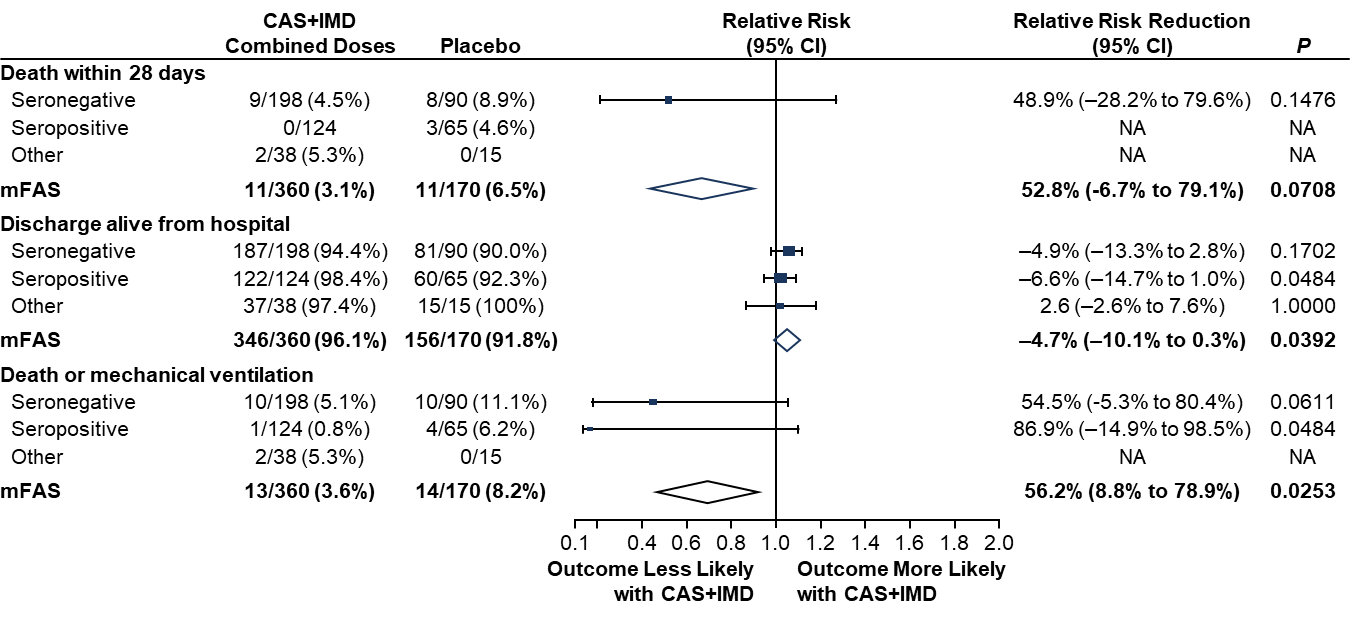

Panel A shows a forest plot of relative risk and relative risk reduction with 95% CI for patients on low-flow oxygen (cohort 1, phase III) combined dose analysis of 2.4 g and 8.0 g CAS+IMD versus placebo. Parameters examined include death within 28 days, discharge alive from hospital, and death or mechanical ventilation. Populations analyzed include patients who tested negative for all SARS-CoV-2 antibodies at baseline (seronegative mFAS) patients who tested positive for any SARS-CoV-2 antibody at baseline (seropositive mFAS), those with inconclusive or missing baseline serology (other), and the overall population regardless of serostatus (overall mFAS). *P* values are considered nominal. Panel B shows a forest plot similar to Panel A, but for patients on no supplemental oxygen (cohort 1A, phase II) combined dose analysis of 2.4 g and 8.0 g CAS+IMD versus placebo.

Abbreviations: CI, confidence interval; mFAS, modified full analysis set; SARS-CoV-2, severe acute respiratory syndrome coronavirus 2.

Supplementary Figure 6. Mortality through study day 57 by serostatus

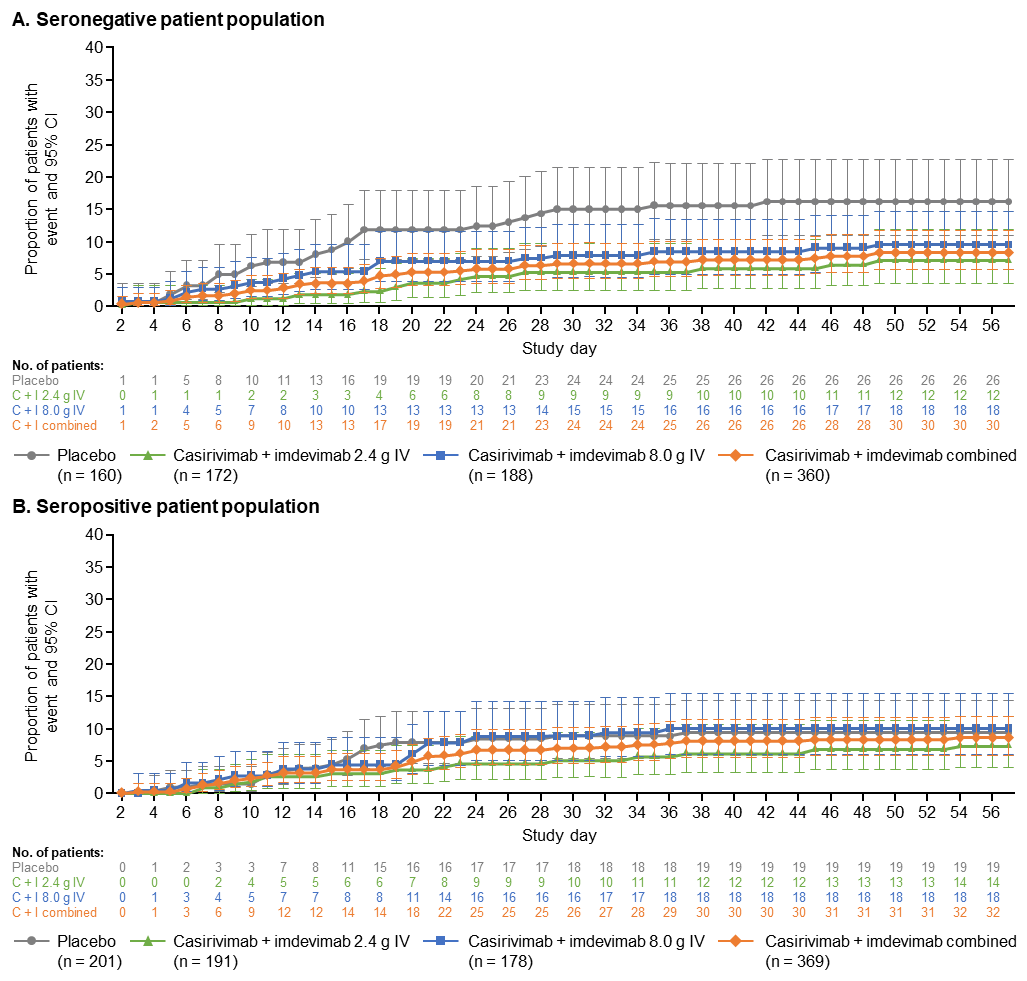

Panel A shows a line plot for the proportion of death in seronegative patients (tested negative for all SARS-CoV-2 antibodies at baseline) who died through study day 57, after administration of CAS+IMD (2.4 g, 8.0 g, or combined analysis of 2.4 g and 8.0 g casirivimab+imdevimab) or placebo. Panel B shows a line plot for the proportion of death similar to panel A but for seropositive patients (tested positive for any SARS-CoV-2 antibodies at baseline).

Abbreviations: IV, intravenous; KM, Kaplan–Meier; SARS-CoV-2, severe acute respiratory syndrome coronavirus 2.
